# Reallocation of 24-hour physical behaviour composition and mortality: exploring effect modification by sleep characteristics

**DOI:** 10.64898/2026.03.23.26349126

**Authors:** Wenxin Bian, Matthew N. Ahmadi, John J. Mitchell, Raaj Kishore Biswas, Nicholas A. Koemel, Dorothea Dumuid, Sebastien F.M. Chastin, Joanna M. Blodgett, Jean-Philippe Chaput, Mark Hamer, Emmanuel Stamatakis

## Abstract

**Background:** Time compositions spent in physical behaviours (sleep, physical activity, posture) is associated with premature mortality, but the moderating role of sleep remains unclear. This study investigates the associations of time reallocations between physical behaviours with all-cause, cardiovascular disease (CVD) and physical activity-related cancer mortality, and the potential effect modification by sleep duration and regularity.

**Methods:** Population-based prospective cohort study of 58,149 adults from the UK Biobank accelerometry subsample, collected between 2013 and 2016, followed up to 2022. Daily sleep duration, sleep regularity index (SRI), sedentary behaviour (SB), standing, light-intensity (LPA) and moderate-to-vigorous physical activity (MVPA) were measured from wrist-worn accelerometers. Participants were categorized based on age-specific sleep duration recommendations, and sample-based sleep regularity index. Associations of behavioural reallocations with all-cause, major CVD, and PA-related cancer mortality were examined using compositional Cox regression.

**Results:** Over a mean follow-up of 8.0 years, 2,209 mortality events occurred (345 CVD and 997 cancer). Displacing sleep with MVPA was associated with lower all-cause and CVD mortality risk in the whole sample, regardless of sleep duration and regularity. Among participants who meet sleep duration guidelines, reallocating 30 minutes from sleep to standing, LPA or MVPA was favourably associated with all-cause mortality with HRs of 0.86 (95%CI 0.79, 0.93), 0.87 (0.80, 0.95), and 0.80 (0.73, 0.87), respectively. Reallocating 30 minutes from sleep to SB, standing, or LPA was adversely associated with CVD risk (HRs 1.08 (1.02, 1.15), 1.10 (1.01, 1.20), and 1.11 (1.03, 1.20)) among those not meeting guidelines. Beneficial associations of reallocating SB to sleep were evident only amongst short (<7h/day) or regular (SRI>87.8) sleepers across all-cause, CVD and cancer mortality.

**Conclusions:** Sleep duration and regularity may modify the theoretical replacement effects of sleep and other physical behaviours on mortality. Replacing sleep with equivalent amounts of MVPA appears largely protective; however, replacing sleep with lower intensity of activity (LPA and standing) is only protective for those who are already meeting sleep duration guidelines. Health effects of increasing sleep may depend on initial sleep regularity and duration. Future personalised behavioural interventions could be optimised by incorporating baseline sleep characteristics in their design and behavioural targets.

## BACKGROUND

Physical activity (PA), sedentary behaviour (SB), sleep and posture are collectively referred to as physical behaviours and are key lifestyle factors that influence health via multiple pathways. Previous studies have shown that poor sleep, excessive sedentary behaviour, and physical inactivity are adversely associated with all-cause, cardiovascular disease (CVD) and cancer mortality risk^1–4^. The national sleep foundation (NSF) guidelines recommend 7-9 hours of sleep per day for adults (aged 18-64 years) and 7-8 for older adults (aged ≥65 years)^5^, while global health guidelines strongly emphasizing PA and advise limiting sedentary time^6,7^. However, because physical behaviours share a finite 24-hour day, increasing time in one behaviour (e.g., sleep) inevitably reduces time spent in others (e.g., PA and/or SB). Previous guidelines do not account for time-reallocation effects and are based on uniform assumptions, overlooking inter-individual variability in behavioural distributions. There is emerging prospective evidence^8,9^ that a change in time composition between behaviour is associated with health improvements.

The compositional nature of interconnected daily physical behaviours implies that no single behaviour alone can determine an individual’s overall health status. Compositional data analysis^10^ is increasingly used to examine the associations of 24-hour behaviour compositions and health outcomes^8,9,11-19^. Population-level evidence supports that reallocating time from SB to light-intensity PA (LPA) and moderate-to-vigorous PA (MVPA) is particularly beneficial for cardiometabolic health^16,19^, lower risk of mortality^8^, incident CVD^13^ and cancer^20^. However, the replacement effects of sleep remain inconsistent and unclear. While beneficial associations have been observed for replacing SB with sleep, and sleep with LPA or MVPA in relation to mortality risk^12,21^, a recent pooled analysis found no significant associations between sleep duration and all-cause mortality^8^. It is well established that healthy sleep characteristics, such as adequate sleep duration and high sleep regularity^22^, play a key regulatory role in normal physiological processes, including glucose homeostasis, inflammation and endocrine function, and are independently associated with improved health outcomes^23,24^. For example, short and long sleep may amplify the inverse association between PA and mortality or cardiometabolic health25,26, while maintaining regular sleep may counteract some of the adverse effects of inadequate sleep^27–29^. Irregular sleep, the inconsistency of sleep-wake time closely linked to circadian rhythm disruption, has been relatively understudied, yet evidence suggests it may be a stronger predictor of mortality risk than short sleep duration alone^30^. These findings suggest that the effects of reallocating time among behaviours may vary depending on an individual’s sleep characteristics.

It is unknown whether behavioural time reallocation associations differ for people with sub-optimal sleep behaviours (very short or long sleep durations, irregular sleep timings). Guidelines often emphasize increasing PA without fully accounting for the influence of sleep. No study to date has examined how sleep duration and regularity may modify associations between 24-hour behavioural reallocations and mortality risk. We hypothesised that participants with short sleep would benefit more from reallocations of lower intensity behaviours to sleep. The aims of this study were to examine how reallocating sleep to and from other behaviours is associated with all-cause, CVD, and cancer mortality; and assess whether sleep duration and regularity modified any such reallocation effects.

## METHODS

### Sample

Participants were drawn from the UK Biobank accelerometry sub-study, in which individuals wore a wrist accelerometer (Axivity AX3) on their dominant wrist 24 hours/day continuously for 7 days between June 2013 and January 2016 to capture daily rest and activity patterns^31,32^. Baseline participant characteristics were measured via questionnaire, interviews, and physical measurements upon enrolment between 2006 and 2010. The ethical approval was received from the UK National Health service (NHS) and National Research Ethics Service for the UK (No. 11/NW/0382). All participants provided written informed consent and were followed up through linkage to their health-related records. Our sample included participants with at least 5 days of valid accelerometer data (≥16h of wear-time with sleep data)^27,28^ including at least 1 day being a weekend day to account for weekday–weekend variation^29,33^. We excluded participants with missing covariate data, prevalent CVD or cancer (depending on the study outcome) at accelerometry assessment, or an event during first year of follow-up.

### Physical behaviours

Physical behaviours were measured from wrist-worn accelerometer. Sleep period time was calculated using a sleep detection algorithm, which has been validated against polysomnography with high accuracy (87%-94%)^34^. A validated random forest activity classifier^35^ with strong overall accuracy of 84.6%^36^ was used to detect time spent in SB (sitting or reclining), standing utilitarian movements, LPA (light ambulatory activities <100mg) and MVPA (moderate to vigorous ambulatory activities ≥100mg and high energetic activities)^1,35,37,38^. Daily time spent in each behaviour was calculated by averaging the time (minutes) across valid wear days. Sleep duration was categorised according to the NSF’s age-specific recommendations: 7–9 hours/day for adults aged 18–64 years, and 7–8 hours/day for those aged ≥65 years^5^. Participants were classified as (1) meeting or (2) not meeting the recommended sleep duration, with those not meeting recommendations further subdivided into (3) short sleepers (less than recommended) and (4) long sleepers (more than recommended). We assessed sleep regularity using the sleep regularity index^22^ (SRI; scaled from 0 to 100) that captures day-to-day variability of sleep timing, including bedtime, waketime, sleep durations, naps and awakenings after sleep onset. Regular sleep (SRI>87.8; top quartile, percentile 75-100%) and irregular sleep (SRI≤87.8; percentile 0-75%) were categorized based on sample SRI distribution and recent research in the UK Biobank^27,31^.

### Covariates

We selected covariates based on previous PA and sleep-specific UK Biobank literature^1,3,28^. These included common demographic and clinical covariates such as age, sex, ethnicity, body mass index (BMI), smoking status, alcohol consumption, fruit and vegeTable consumption, tea and coffee intake (major contributors of caffeine in the diet), education, self-reported sleep problems (including insomnia symptoms, daytime sleepiness, snoring), shift work status, medication use (cholesterol, diabetes, blood pressure), and family history of CVD or cancer. Full covariate definitions are provided in **eTable 1**.

### Outcomes

Mortality data were obtained through data linkage with the National Health Service (NHS) Digital of England and Wales and the NHS Central Register and National Records of Scotland followed up to 30 November 2022. Inpatient hospitalization data were sourced from the Hospital Episode Statistics for England, Scottish Morbidity Record and Patient Episode Database for Wales. Cancer diagnosis data were linked through national cancer registries, with follow-up available until 31 December 2020 for England, 31 December 2016 for Wales, and 30 November 2021 for Scotland. We identified major CVD and PA-related cancer diseases using ICD-10 (International Classification of Diseases) codes with full details provided in **eTable 2**.

### Statistical analysis

We winsorised the range of exposure values at the 2.5^th^ and 97.5^th^ percentile for sleep, SB, standing and LPA to limit the potential influence of sparse data or outliers. As the 2.5^th^ percentile was 0 for MVPA, we capped the data at the 97.5^th^ percentile. The log-ratio EM algorithm^39^ was applied for the estimation of rounded zeros^40^. We used compositional Cox regression to investigate associations between behaviour compositions and all-cause, CVD and cancer mortality risk. A composition was described as the proportional allocation of daily time across the five physical behaviours. Daily behaviour compositions in each event group were compared to the sample average using a multivariate analysis of variance Pillai’s trace test. Isometric log-ratio (<u>ilr</u>) pivot coordinates were applied to model the relative time spent across the various behaviours in relation to each outcome^10,16-18^. All possible pivot coordinate permutation of the behaviours were fitted, and the first coordinate of each model was reported in **eTable 3**. Schoenfeld residuals were used to test the proportional hazards assumptions and were satisfied. We performed minute-by-minute isotemporal substitution to estimate risk associated with reallocating time between sleep and movement behaviours (e.g., replacing 30min of sleep with 30min of SB), around the mean composition while holding other behaviours constant^16,18^. Statistical interaction tests were performed prior to the stratified analyses (**eTable 4**). Analyses were stratified by the predefined sleep duration and regularity groups, adjusted for all covariates, and additionally for SRI in the sleep duration models.

We conducted sensitivity analyses excluding participants with (i) events within the first two years of follow-up, (ii) medication use for cholesterol, diabetes or blood pressure, (iii) self-reported poor sleep characteristics, including insomnia symptoms, daytime sleepiness, snoring (≤1 sleep score^3^), (iv) prevalent CVD or cancer at accelerometry baseline for all-cause mortality; we also additionally adjusted for seasons of data collection, applied more stringent categorization of sleep regularity (bottom quintile, SRI≤70.6 vs top quintile, SRI>89.0)^22^, and conducted additional analyses for all-cause cancer mortality given the primary focus on PA-related cases. All analyses were conducted in R version 4.4.1, using the *Compositions* and *zCompositions* functions.

## RESULTS

### Sample and events

The analytic sample included 58,149 participants, with 2,209 deaths over a mean (SD) 8.0 (0.9) years of follow-up (345 CVD, 997 PA-related cancer; details in **eFigure 1**). Participants had a mean age of 62 (7.8) years and 56.1% were female, with a compositional mean of 7h 42min sleep, 10h 42min SB, 2h 51min standing, 1h 53min LPA, and 33min MVPA (**eFigure 2**). Log-ratio differences in compositional means by mortality status showed that individuals who died spent 4.5% more time sedentary and 1.2%, 5.1%, 4.6%, and 37.7% less time in sleep, standing, LPA, and MVPA, respectively (**eFigure 3**). Baseline sample characteristics by sleep duration (meeting; not meeting recommendations) and regularity (regular; irregular) are shown in **Table 1** and **eTable 5**.

**Table 1:**
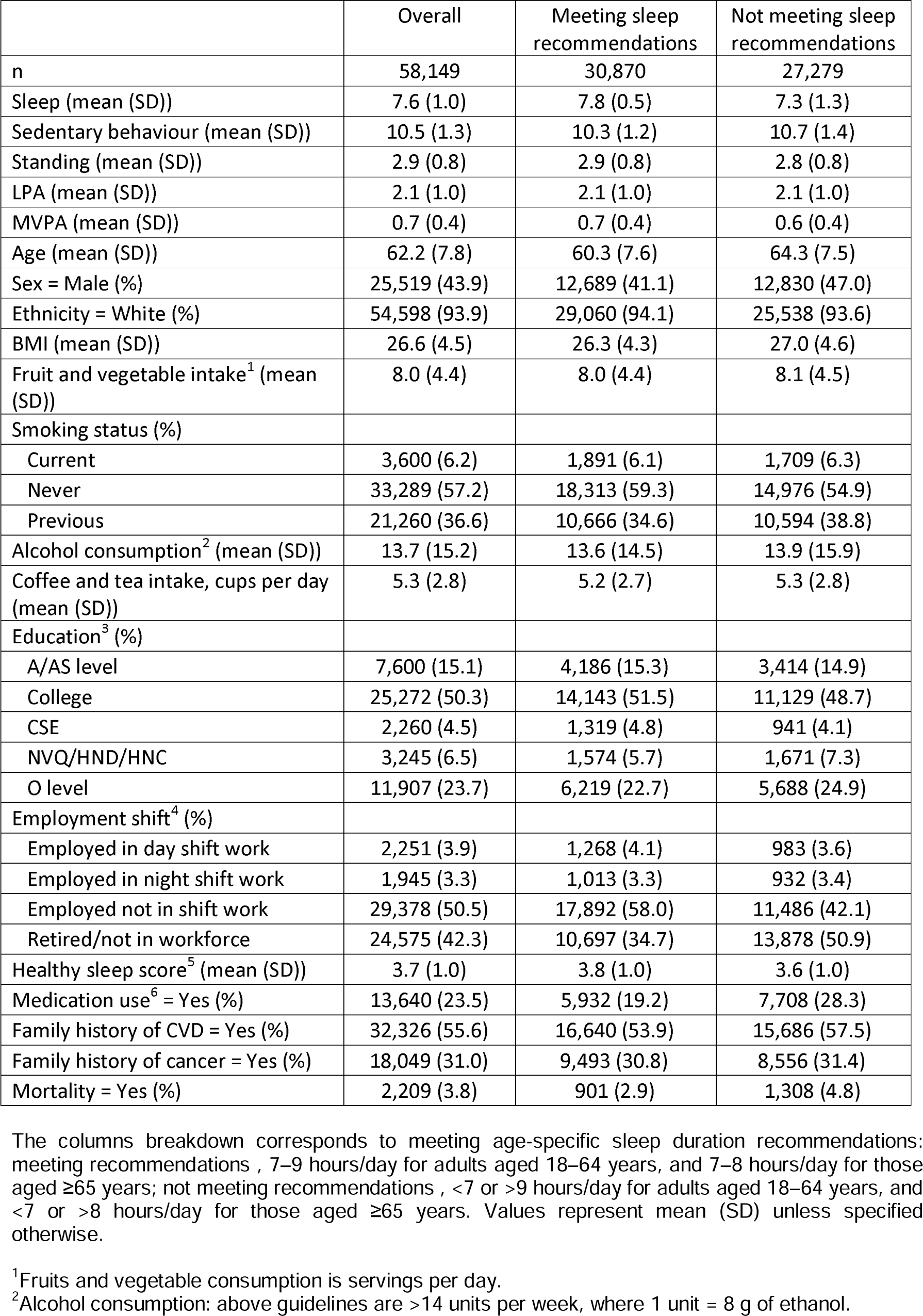
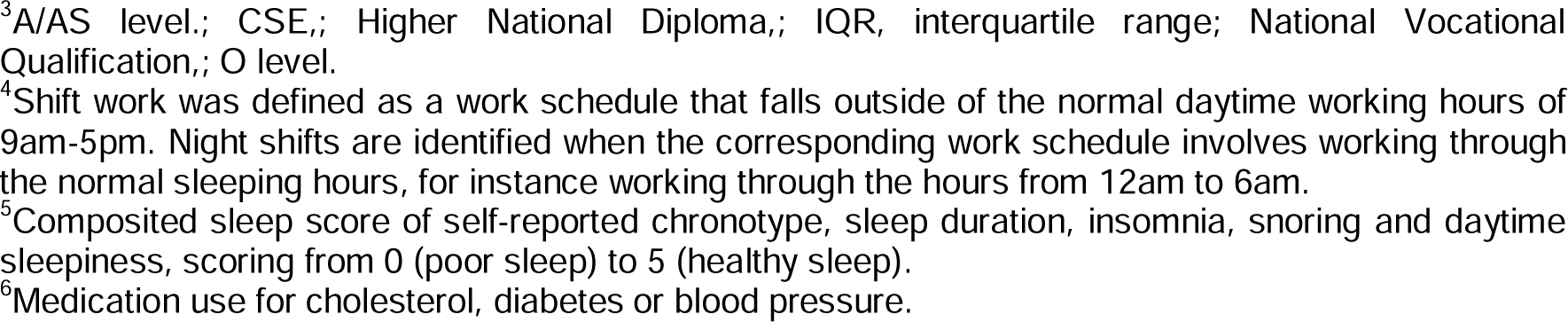
Baseline characteristics of participants stratified by participants whether meeting sleep duration guidelines group (n=58,149).

|  | Overall | Meeting sleep recommendations | Not meeting sleep recommendations |
| --- | --- | --- | --- |
| n | 58,149 | 30,870 | 27,279 |
| Sleep (mean (SD)) | 7.6 (1.0) | 7.8 (0.5) | 7.3 (1.3) |
| Sedentary behaviour (mean (SD)) | 10.5 (1.3) | 10.3 (1.2) | 10.7 (1.4) |
| Standing (mean (SD)) | 2.9 (0.8) | 2.9 (0.8) | 2.8 (0.8) |
| LPA (mean (SD)) | 2.1 (1.0) | 2.1 (1.0) | 2.1 (1.0) |
| MVPA (mean (SD)) | 0.7 (0.4) | 0.7 (0.4) | 0.6 (0.4) |
| Age (mean (SD)) | 62.2 (7.8) | 60.3 (7.6) | 64.3 (7.5) |
| Sex = Male (%) | 25,519 (43.9) | 12,689 (41.1) | 12,830 (47.0) |
| Ethnicity = White (%) | 54,598 (93.9) | 29,060 (94.1) | 25,538 (93.6) |
| BMI (mean (SD)) | 26.6 (4.5) | 26.3 (4.3) | 27.0 (4.6) |
| Fruit and vegetable intake<sup>1</sup> (mean (SD)) | 8.0 (4.4) | 8.0 (4.4) | 8.1 (4.5) |
| Smoking status (%) |  |  |  |
| Current | 3,600 (6.2) | 1,891 (6.1) | 1,709 (6.3) |
| Never | 33,289 (57.2) | 18,313 (59.3) | 14,976 (54.9) |
| Previous | 21,260 (36.6) | 10,666 (34.6) | 10,594 (38.8) |
| Alcohol consumption<sup>2</sup> (mean (SD)) | 13.7 (15.2) | 13.6 (14.5) | 13.9 (15.9) |
| Coffee and tea intake, cups per day (mean (SD)) | 5.3 (2.8) | 5.2 (2.7) | 5.3 (2.8) |
| Education<sup>3</sup> (%) |  |  |  |
| A/AS level | 7,600 (15.1) | 4,186 (15.3) | 3,414 (14.9) |
| College | 25,272 (50.3) | 14,143 (51.5) | 11,129 (48.7) |
| CSE | 2,260 (4.5) | 1,319 (4.8) | 941 (4.1) |
| NVQ/HND/HNC | 3,245 (6.5) | 1,574 (5.7) | 1,671 (7.3) |
| O level | 11,907 (23.7) | 6,219 (22.7) | 5,688 (24.9) |
| Employment shift<sup>4</sup> (%) |  |  |  |
| Employed in day shift work | 2,251 (3.9) | 1,268 (4.1) | 983 (3.6) |
| Employed in night shift work | 1,945 (3.3) | 1,013 (3.3) | 932 (3.4) |
| Employed not in shift work | 29,378 (50.5) | 17,892 (58.0) | 11,486 (42.1) |
| Retired/not in workforce | 24,575 (42.3) | 10,697 (34.7) | 13,878 (50.9) |
| Healthy sleep score<sup>5</sup> (mean (SD)) | 3.7 (1.0) | 3.8 (1.0) | 3.6 (1.0) |
| Medication use<sup>6</sup> = Yes (%) | 13,640 (23.5) | 5,932 (19.2) | 7,708 (28.3) |
| Family history of CVD = Yes (%) | 32,326 (55.6) | 16,640 (53.9) | 15,686 (57.5) |
| Family history of cancer = Yes (%) | 18,049 (31.0) | 9,493 (30.8) | 8,556 (31.4) |
| Mortality = Yes (%) | 2,209 (3.8) | 901 (2.9) | 1,308 (4.8) |

The columns breakdown corresponds to meeting age-specific sleep duration recommendations: meeting recommendations , 7–9 hours/day for adults aged 18–64 years, and 7–8 hours/day for those aged ≥65 years; not meeting recommendations , <7 or >9 hours/day for adults aged 18–64 years, and <7 or >8 hours/day for those aged ≥65 years. Values represent mean (SD) unless specified otherwise.
<sup>1</sup>Fruits and vegetable consumption is servings per day.
<sup>2</sup>Alcohol consumption: above guidelines are >14 units per week, where 1 unit = 8 g of ethanol.
<sup>3</sup>A/AS level.; CSE.; Higher National Diploma.; IQR, interquartile range; National Vocational Qualification.; O level. <sup>4</sup>Shift work was defined as a work schedule that falls outside of the normal daytime working hours of 9am-5pm. Night shifts are identified when the corresponding work schedule involves working through the normal sleeping hours, for instance working through the hours from 12am to 6am. <sup>5</sup>Composited sleep score of self-reported chronotype, sleep duration, insomnia, snoring and daytime sleepiness, scoring from 0 (poor sleep) to 5 (healthy sleep). <sup>6</sup>Medication use for cholesterol, diabetes or blood pressure.

### Overall associations of physical behaviours with all-cause, CVD and cancer mortality

Using multivariable-adjusted substitution models, reallocation between sleep and MVPA showed the strongest and consistent associations with mortality outcomes, compared to sleep vs. other behavioral substitutions (**eFigure 4**). For instance, reallocating 30 minutes of sleep to MVPA was associated with lower all-cause, CVD and cancer mortality risks, with corresponding HRs of 0.82 (95%CI 0.79, 0.86), 0.82 (0.75, 0.90), and 0.92 (0.87, 0.98). Replacing sleep with SB or LPA was associated with higher CVD mortality risk, while replacing SB with sleep was protective for all-cause and CVD mortality. Reallocations between sleep and standing or LPA showed no statistically significant replacement effects on any outcomes. Substitution models of all other behaviours are presented in **eFigure 5-7**.

### Associations of physical behaviours with all-cause mortality by sleep duration

We identified distinct dose-response associations between time reallocation (sleep vs. movement behaviours) and all-cause mortality among participants with different sleep characteristics (**Figure 1**). For participants meeting sleep duration recommendations, reallocating time from sleep to standing, LPA, or MVPA was associated with lower mortality risk, while the reverse reallocations were associated with higher risks (**Figure 1A**). Beneficial associations of reallocating time from sleep to MVPA were most pronounced, followed by LPA and standing. For example, reallocating 30 minutes of sleep to standing, LPA or MVPA was associated with lower HRs corresponding to 0.86 (95% CI 0.79, 0.93), 0.87 (0.80, 0.95), and 0.80 (0.73, 0.87). Associations were attenuated among participants not meeting recommendations (except for MVPA) and were largely driven by short sleepers (**Figure 1B, 1C**). Among short sleepers, reallocating time (e.g., 30–60 minutes) from LPA to sleep was associated with HRs of 0.92 (0.85, 0.99)–0.82 (0.70, 0.97), while reallocating same amount from sleep to LPA was associated with HRs of 1.08 (1.01, 1.16)–1.16 (1.01, 1.34) (**Figure 1C**). For long sleepers, replacing sleep with MVPA was most strongly associated with lower mortality risk (**Figure 1D**). Substitution models of all other behaviours are presented in **eFigure 8**.

**Figure 1.**
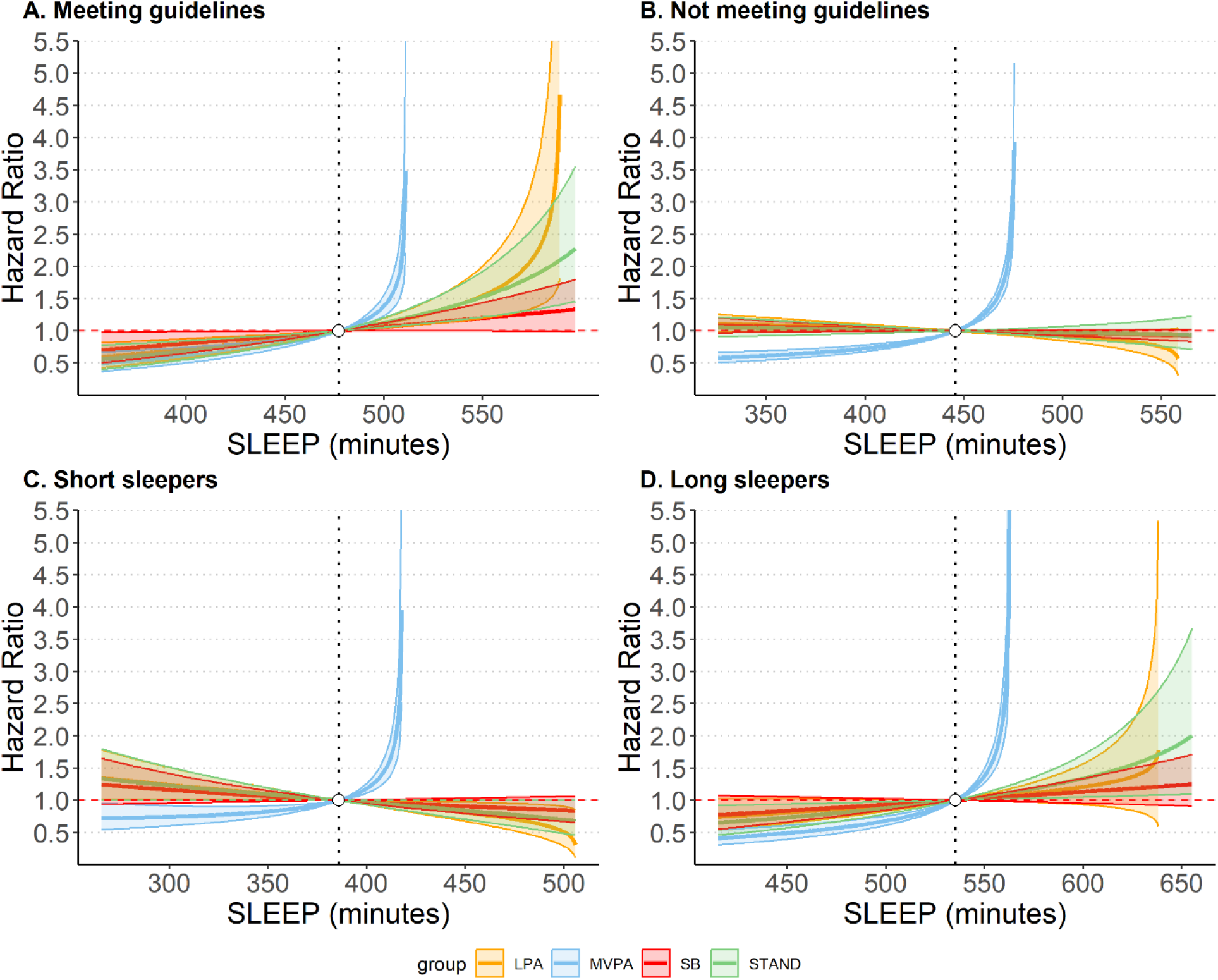
Associations of time reallocation between sleep and other movement behaviours with all-cause mortality stratified by sleep duration (n=58149, 2209 events). Estimated hazard ratios with 95%CIs in all-cause mortality associated with the reallocation of time between sleep and other behaviours across four sleep duration groups: A. meeting sleep duration recommendations (7– 9 hours/day for ages 18–64 and 7–8 hours/day for ages ≥65) (n=30,870; 901 events); B. not meeting recommendations (n=27,279; 1,308 events); C. short sleep (below recommendations) (n=15,739; 711 events); D. long sleep (above recommendations) (n=11,540; 597 events). Reference (dashed vertical line) is the sample compositional mean sleep duration: A. 477min (7.95h), B. 446min (7.43h), C. 386min (6.43h), D. 535min (8.92h). To the left of the reference shows replacing sleep with other behaviours, while to the right shows replacing other behaviours with sleep. Models adjusted for age, sex, ethnicity, SRI, BMI, diet, smoking, alcohol consumption, tea and coffee consumption, education, employment shift, sleep problems, medications, parental history of CVD, parental history of cancer, prevalent CVD and prevalent cancer.

### Associations of physical behaviours with CVD mortality by sleep duration

Substitution models for CVD mortality by sleep duration revealed patterns similar to all-cause mortality but with wider 95%CIs (**Figure 2, eFigure 9**). Replacing sleep with MVPA was associated with lower CVD mortality risk in both groups, while replacing sleep with SB or LPA was associated with higher risks among those not meeting recommendations. For example, relative to the compositional mean of 7.4 hours, reallocating 30 minutes from sleep to MVPA was associated with a HR of 0.75 (0.62, 0.92) for those meeting sleep duration recommendations and 0.78 (0.70, 0.87) for those not (**Figure 2A, 2B**). For those not meeting recommendations, reallocating 30 minutes of sleep to SB, standing, or LPA was associated with higher HRs corresponding to 1.08 (1.02, 1.15), 1.10 (1.01, 1.20), and 1.11 (1.03, 1.20). Reallocating time (e.g., 60 minutes) from SB or LPA to sleep was associated with lower CVD mortality risks (HRs 0.86 (0.77, 0.96) and 0.76 (0.61, 0.95), respectively). Results for short and long sleepers remained similar (**Figure 2C, 2D**).

**Figure 2.**
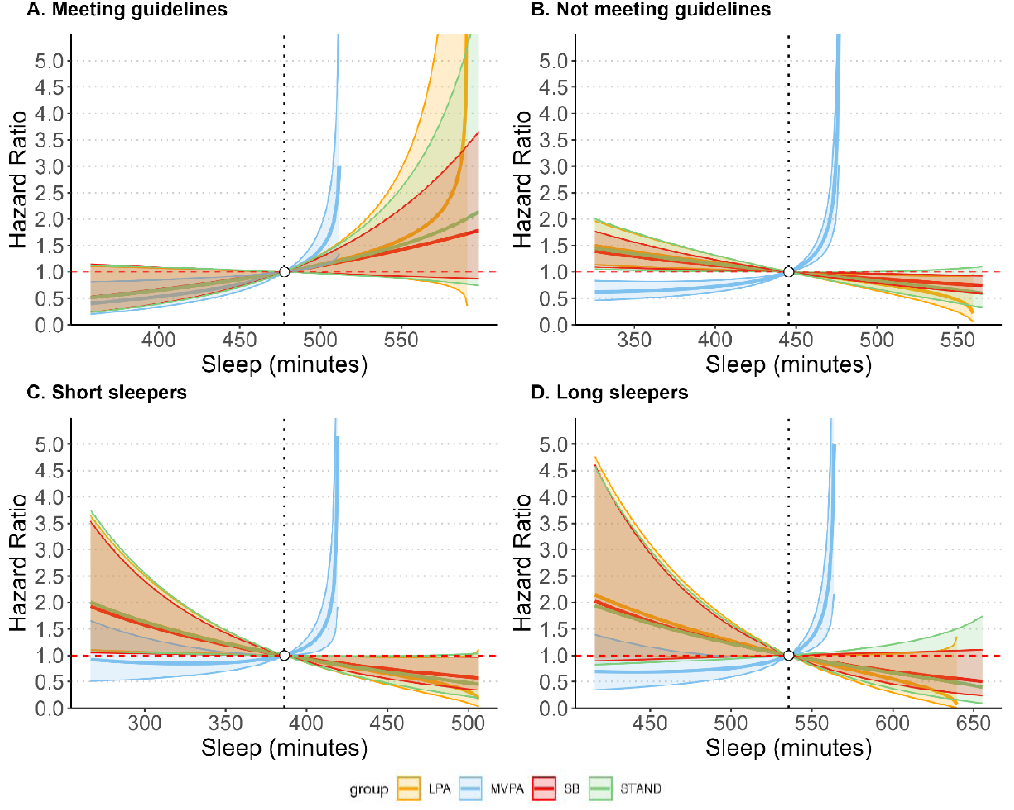
Associations of time reallocation between sleep and other movement behaviours with CVD mortality stratified by sleep duration (n=53025, 435 events). Estimated hazard ratios with 95%CIs in CVD mortality associated with the reallocation of time between sleep and other behaviours across four sleep duration groups: A. meeting sleep duration recommendations (7–9 hours/day for ages 18–64 and 7–8 hours/day for ages ≥65) (n=28,653; 169 events); B. not meeting recommendations (n=24,372; 266 events); C. short sleep (below recommendations) (n=14,184; 156 events); D. long sleep (above recommendations) (n=10,188; 110 events). Reference (dashed vertical line) is the sample compositional mean sleep duration: A. 478min (7.97h), B. 445min (7.42h), C. 386min (6.43h), D. 536min (8.93h). To the left of the reference shows replacing sleep with other behaviours, while to the right shows replacing other behaviours with sleep. Models adjusted for age, sex, ethnicity, SRI, BMI, diet, smoking, alcohol consumption, tea and coffee consumption, education, employment shift, sleep problems, medications and parental history of CVD.

### Associations of physical behaviours with cancer mortality by sleep duration

Results for cancer mortality were similar to above such that among participants meeting sleep duration recommendations, reallocating time from sleep and SB to standing and LPA had protective associations, whereas reallocating time in the opposite direction had adverse associations (**Figure 3A, eFigure 10A**). For example, reallocating 30 min from sleep to standing and LPA was associated with 12% lower cancer mortality risks with corresponding HRs of 0.88 (0.78, 0.99) and 0.88 (0.79, 0.99), whereas reallocating 30 min to sleep was associated with higher risks with corresponding HRs of 1.16 (1.02, 1.31) and 1.16 (1.02, 1.31), respectively. Among those not meeting sleep duration recommendations, no significant association was observed for the time reallocations between lower intensity behaviors (sleep, SB, standing, and LPA) in either direction (**Figure 3B-D, eFigure 10B**).

**Figure 3.**
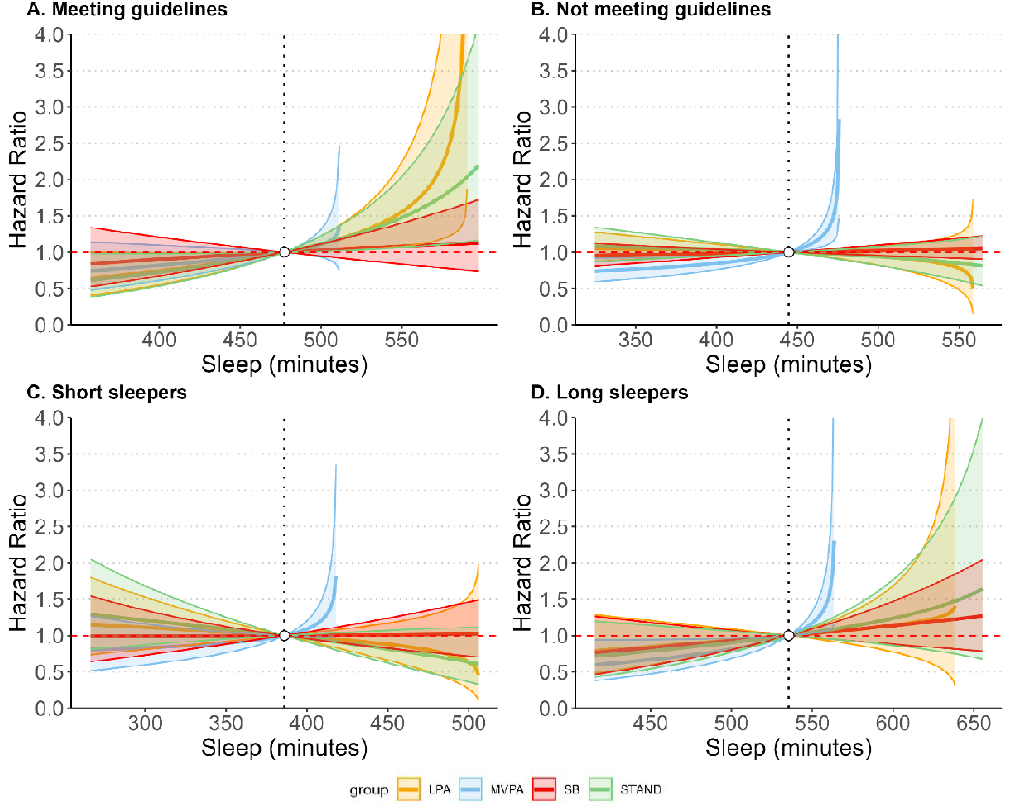
Associations of time reallocation between sleep and other movement behaviours with cancer mortality stratified by sleep duration (n=53209, 997 events). Estimated hazard ratios with 95%CIs in cancer mortality associated with the reallocation of time between sleep and other behaviours across four sleep duration groups: A. meeting sleep duration recommendations (7–9 hours/day for ages 18–64 and 7–8 hours/day for ages ≥65) (n=28,584; 435 events); B. not meeting recommendations (n=24,625; 562 events); C. short sleep (below recommendations) (n=14,415; 298 events); D. long sleep (above recommendations) (n=10,210; 264 events). Reference (dashed vertical line) is the sample compositional mean sleep duration: A. 477min (7.95h), B. 445min (7.41h), C. 386min (6.43h), D. 535min (8.92h). To the left of the reference shows replacing sleep with other behaviours, while to the right shows replacing other behaviours with sleep. Models adjusted for age, sex, ethnicity, SRI, BMI, diet, smoking, alcohol consumption, tea and coffee consumption, education, employment shift, sleep problems, medications and parental history of cancer.

### Associations of physical behaviours with mortality by sleep regularity

Relative to sleep duration, dose-response associations of sleep reallocation with mortality outcomes were less evident when comparing regular and irregular sleepers (**Figure 4, eFigure 11, 12**). Replacing MVPA with sleep or sleep with SB was adversely associated with all-cause mortality, while the reverse displacement was favourably associated. The protective associations of more MVPA among irregular sleepers were more robust compared to regular sleepers. For example, reallocating 30 min from sleep to MVPA was associated with 14% (HR [95%CI] 0.86 [0.79, 0.94]) and 18% (0.82 [0.78, 0.85]) lower mortality risks among regular and irregular sleepers, respectively. Among regular sleepers, reallocating 30 min from SB to sleep was associated with 7% (0.93 [0.88, 0.98]) lower risks, whereas the reverse reallocation was associated with 7% (1.07 [1.02, 1.14)) higher risks. There was no such association among irregular sleepers. No association was found between reallocations of sleep with standing or LPA in both groups. Similar associations were observed for CVD and cancer mortality (**eFigure 11**).

**Figure 4.**
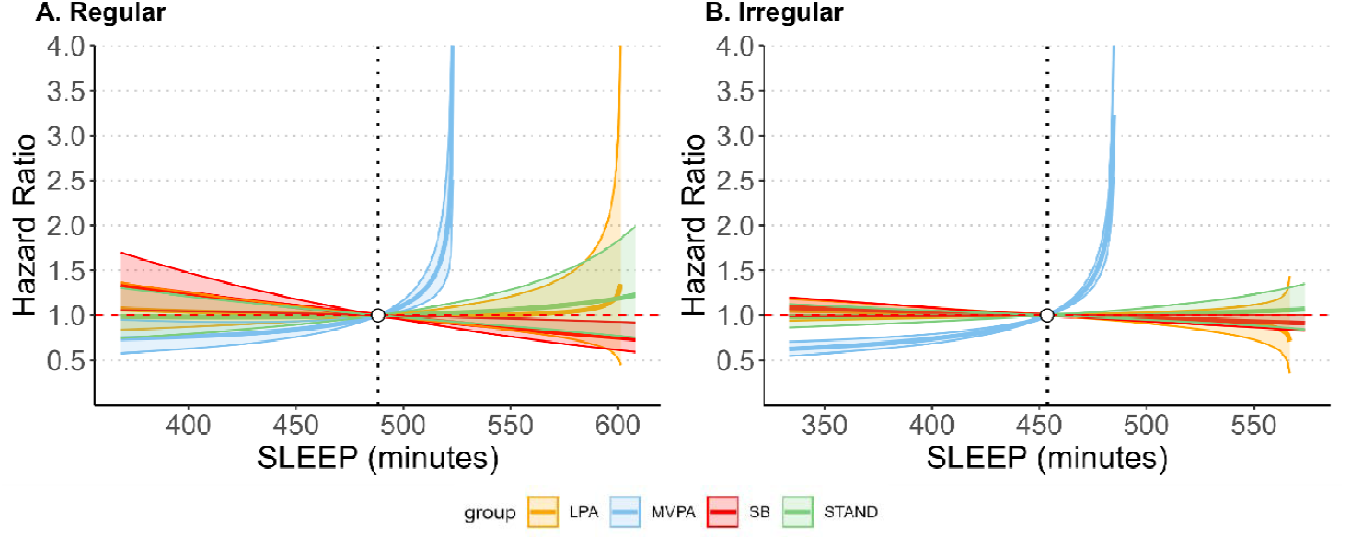
Associations of time reallocation between sleep and other movement behaviours with all-cause mortality stratified by sleep regularity (n=58149, 2209 events). Estimated hazard ratios with 95%CIs in all-cause mortality associated with the reallocation of time between sleep and other behaviours across sleep regularity groups: A. regular sleep (n=14,507; 517 events); B. irregular sleep (n=43,642; 1,692 events). Reference (dashed vertical line) is the sample compositional mean sleep duration: A. 488min (8.13h), B. 454min (7.56h). To the left of the reference shows replacing sleep with other behaviours, while to the right shows replacing other behaviours with sleep. Models adjusted for age, sex, ethnicity, BMI, diet, smoking, alcohol consumption, tea and coffee consumption, education, employment shift, sleep problems, medications, parental history of CVD, parental history of cancer, prevalent CVD and prevalent cancer.

### Sensitivity analyses

Results remained consistent when excluding participants with mortality within the first two years of follow-up (**eFigure 13-16**), poor sleep characteristics (**eFigure 17-20**); adjusting for seasonality (**eFigure 21-24**); applying a stricter sleep regularity classification (**eFigure 25**); and substituting all-cause for PA-related cancer mortality (**eFigure 26**). Associations for reallocations between sleep and low-intensity behaviours were attenuated after excluding participants on medication for cholesterol, diabetes or blood pressure (**eFigure 27-30**), and excluding prevalent CVD and cancer for all-cause mortality (**eFigure 31**), possibly due to reduced statistical power.

## DISCUSSION

We examined the prospective associations between device-based measures of 24-hour physical behaviours with all-cause, CVD and cancer mortality stratified by sleep duration recommendations and sleep regularity characteristics. This is the first study to investigate whether sleep characteristics (duration and regularity) modify the associations between time reallocation of 24-hour physical behaviours and prospective outcomes. We found that reallocating time from sleep to MVPA was favourably associated with mortality outcomes. Beneficial associations of reallocating sleep to standing or LPA with all-cause mortality were observed only among participants already meeting or exceeding sleep recommendations. Protective associations of reallocating SB to sleep with mortality outcomes were observed only in regular sleepers. Our findings indicate that reallocating sleep beyond the minimum recommendation to PA may yield health benefits, with MVPA being the most beneficial, followed by LPA, and standing. The absence of mortality benefits from increasing sleep duration among irregular sleepers may reflect the dominant influence of disrupted circadian patterns on health outcomes.

Our study contributes novel insights to the literature on time reallocation of daily physical behaviours and mortality risk. In addition to previous compositional substitution findings that have reported either null^41^ or beneficial effects^9,15,21,42^ on all-cause mortality displacing sleep with low intensity physical activity, our results suggest that such modelled effects vary considerably by sleep duration status. Prior studies reported beneficial association with mortality when displacing sleep to LPA^9^, whereas extending sleep at the expense of other behaviours was associated with increased major adverse cardiac events and mortality risk, though the increase was marginal when replacing SB or LPA^42^. Previous work^21^ indicated that insufficient sleep duration should be addressed by reallocating time from SB but not from LPA or MVPA. Using stratified approach, our study showed that the beneficial association of replacing sleep with low intensity behaviours was evident only among individuals with sufficient sleep but not those without. For short sleepers (<7h/day), reallocating 30-60 minutes from LPA to sleep was associated with 8-18% lower mortality risk, whereas reallocating same amount to LPA was associated with 8-16% higher risk. Conversely, among those meeting or exceeding the recommended sleep durations, reallocating any time to sleep at the expense of MVPA, LPA or standing was adversely associated with mortality. Our findings corroborate those that showed favourable associations of replacing one hour of sleep with PA behaviours only among those who self-reported more than 7 hours of sleep^43^. It is also supported by previous joint associations revealing that the combinations of a low volume of PA with short and long sleep duration were associated with the highest risk of all-cause mortality, while lowest risk of death was observed in the group with normal sleep duration and a high volume of PA^25^.

Distinct associations observed across sleep groups highlight the potential modifying role of sleep. It may reflect underlying differences in restorative sleep, such that when restorative sleep thresholds are not met, the marginal cardiometabolic benefits of low intensity physical activity are insufficient to offset the regular need for resting. Specifically, sleep is essential for physiological recovery, and its deprivation disrupts immune and metabolic functions by impairing blood pressure regulation, increasing inflammation, weakening immune defence, and inducing metabolic imbalances^44,45^. Evidence suggests that reallocating time from SB and LPA to sleep is associated with improved cardiometabolic health markers, such as lower blood pressure^19^, waist circumference, triglyceride and HbA1c^16^, as well as CVD risk^13^. This aligns with our finding where displacing time from SB and LPA to sleep among participants not meeting sleep duration recommendations showed favourable associations with CVD mortality risks. Moreover, Holtermann et al. proposed the “Sweet-spot Hypothesis” suggesting that excessive physical activity without adequate sleep can negatively impact cardiometabolic health and the “sit less, move more” message may not be ideal for all individuals^46^. In line with the established restorative role of sufficient sleep in cardiovascular health, our findings suggest that repayment of sleep debt may be prioritized over the relatively modest benefits of low-intensity activities. However, once minimum restorative sleep thresholds are achieved, benefits gained from physical activity may outweigh those of additional sleep. This appears to align with the widely reported U-shaped association of sleep duration and health outcomes^23^. It is worth noting that the theoretical benefits of replacing sleep with MVPA on all-cause and CVD mortality risk were most prominent^12,15,42^ and remained consistent regardless of sleep status. This may be supported by a recent device-based UK Biobank study showing a stronger overall effect of MVPA on mortality risk compared to sleep, with just an additional 2 min/day of MVPA (relative to the 5^th^ percentile) associated with 10% lower mortality risk, compared to 24 min/day of additional sleep for the same effect^33^.

Our study advances previous literature by elucidating potential moderating role of sleep regularity in time reallocation effects. Our findings demonstrate that sleep regularity may modify the health effects of sleep time reallocation, with irregular sleep potentially overriding the benefits of increased sleep duration. Mortality benefits from reallocating SB to sleep were evident among regular but not irregular sleepers, suggesting that sleep irregularity itself may be a dominant risk factor, potentially negating the benefits of increased sleep duration. This aligns with recent studies linking irregular sleep with major adverse cardiovascular events, type 2 diabetes and mortality, possibly outweighing the relevance of sufficient sleep duration^27,28,30^. It may be because sleep regularity serves as a more direct behavioural indicator of circadian misalignment than sleep duration^30^. Irregular sleep patterns can disrupt the body’s circadian rhythms, leading to hormonal dysregulation and increased insulin resistance^24,45^, regular sleep may provide the circadian stability needed for restorative processes to occur efficiently. Device-based joint analyses support its dominant and independent role, showing consistent cardiometabolic patterns across duration groups^47^. Our study further emphasizes the importance of sleep regularity beyond sleep duration for cardiovascular and overall health.

These findings underscore the importance of integrated public health recommendations and tailored interventions for incorporating sleep regularity and balancing sleep duration and physical activity in mitigating mortality risk. This may include promoting regular sleep beyond sufficient sleep duration; more moderate-to-vigorous activities as priority for adults who already have adequate sleep, while recognizing that light-intensity activities can still be advantageous if higher intensities are not feasible. Conversely, increasing sleep should be prioritised for individuals not meeting minimum sleep recommendations, while encouraging high intensity activities for those exceeding recommended sleep.

### Strength and limitation

Key strengths of our study include the use of a validated^48^ algorithms differentiating between standing and ambulatory LPA. Moreover, the use of compositional data analysis method to modelling time-use data^10^ and performance of stratified analyses, provided a more nuanced understanding of how sleep may affect mortality risk.

Our study also had limitations, including its observational nature which may be subject to potential biases. Our results of behaviour reallocations were theoretical, i.e. based on statistical modelling across participants rather than actual replacements, and within-participant changes cannot be directly addressed. Reverse causality, particularly for sleep duration given the relatively short follow-up, cannot be ruled out. There was a 5.5-year gap between the baseline and accelerometry data collection; however, most covariates remained stable over time, except for employment status and medication^49^. Finally, while the UK Biobank response rate was relatively low (5.5%^50^), it has been shown that associations between health related behaviours and mortality outcomes are not materially compromised by poor representativeness^51^.

## CONCLUSION

Reallocation of 24-hour physical behaviours was associated with mortality risks and may be modified by sleep duration and regularity, with notable differences depending on adherence to sleep duration guidelines. Reallocating time from sleep to MVPA had protective association across the whole sample, regardless of sleep duration and regularity. However, reallocating time from sleep to lower intensity activities (LPA and static postures) may only be protective for those already meeting sleep duration guidelines. Behavioural allocations that prioritise adequate sleep alongside greater MVPA may be most beneficial for individuals with insufficient sleep. Mortality benefits from increased sleep were evident among regular but not irregular sleepers, suggesting an important role of sleep regularity in behavioural risk profiles. Future personalised behavioural interventions could be enhanced by integrating sleep characteristics in their design and targeted behaviours.

## Acknowledgement

This research has been conducted using the UK Biobank resource under application number 25813. The authors would like to thank all the participants and professionals contributing to the UK Biobank. This research used data assets made available by National Safe Haven as part of the Data and Connectivity National Core Study, led by Health Data Research UK in partnership with the Office for National Statistics and funded by UK Research and Innovation. This work uses data provided by patients and collected by the NHS as part of their care and support.

## Ethic approval and consent to participate

The ethical approval was received from the UK National Health service (NHS) and National Research Ethics Service for the UK (No. 11/NW/0382) and participants provided written informed consent. All information and materials in the manuscript are original and have not been submitted for publication elsewhere.

## Data Availability

The data that support the findings of this study are available from the UK Biobank, but restrictions apply to the availability of these data, which were used under license for the current study, and so are not publicly available. Data are however available from the authors upon reasonable request and with the permission of the UK Biobank.

### Disclosures

ES is a paid consultant and holds equity in Complement One, a US-based commercial entity whose products and services relate to heathy lifestyle behaviours. All other authors disclose no conflict of interest for this work.

