## Supplemental material for "Reallocation of 24-hour physical behaviour composition and mortality: exploring effect modification by sleep characteristics"

### eTable 1 Covariate definitions

| **Variable** | **Definition** | **UK Biobank field ID (if applicable)** |
| --- | --- | --- |
| Age | Continuous | 34, 52, accelerometer date timestamp |
| Sex | Female/Male | 31 |
| Ethnicity | White/Others | 21000 |
| Body mass index | Continuous | 23104 |
| Smoking status | Never, previous, current | 20116 |
| Alcohol consumption* | Continuous (units/week) | 1558, 1568, 1578, 1588, 1598, 1608, 5364. 20117 |
| Fruit and vegetable consumption | Continuous (servings/day) | 1309, 1319, 1289, 1299 |
| Tea and coffee intake | Continuous (cups/day) | 1488, 1498 |
| Education | College/University; A/AS level; O levels; CSE; NVQ/HND/HNC; other | 6138 |
| Self-reported sleep problems^#^ | Healthy sleep score (1-5)^1^ | 1200, 1210, 1220 |
| Shift work status | Retired/not in the workforce; Employed not in shift work; Employed in night shift work; Employed in day shift work | 6142, 826, 3426 |
| Use of CVD medication (cholesterol, blood pressure and diabetes) | Yes/No | 6177, 6153 |
| Family history of CVD | Self-reported mother or father diagnosed with heart disease or stroke | 20107, 20110 |
| Family history of Cancer | Self-reported mother or father diagnosed cancer | 20107, 20110 |

^*^The level of overall alcohol consumption as the number of UK units of alcohol (10 mL/unit) consumed per week.

^#^Self-reported sleep problems including Insomnia symptoms, daytime sleepiness, snoring.

### eTable 2 Assessment of diseases used in the study

| **Variable** | **ICD-10** |
| --- | --- |
| Major CVD^1^ | Diseases of the circulatory system, excluding hypertension, diseases of arteries, and lymph. The ICD-10 codes included were: I0, I11, I13, I20-I51, I60-I69. |
| PA-related Cancer^2^ | The definition of total cancer excluded in situ, benign, uncertain, non-melanoma skin cancer, or non-well-defined cancers. The ICD-10 codes included were: C15, C220, C221, C34, C649, C659, C160, C54, C559, C92, C900, C18, C260, C0, C11, C12, C13, C14, C30, C31, C32, C33, C34, C38, C390, C398, C399, C199, C209, C67, C50. |

### eTable 3 Estimated regression coefficients of isometric log ratio (ILR of daily time composition for SB, Sleep, Standing, LPA and MVPA across all four sleep stratums and three outcomes)

| **Stratum** | **Sample size** | **Sleep** | **SB** | **Standing** | **LIPA** | **MVPA** |
| --- | --- | --- | --- | --- | --- | --- |
| **Model 1: All-cause mortality** | | | | | | |
| Meeting sleep guidelines | 30,870 | 1.27(0.38, 2.16)* | -0.17(-0.84, 0.51) | -0.38(-0.78, 0.01) | -0.09(-0.31, 0.13) | -0.44(-0.55, -0.33)* |
| Not meeting sleep guidelines | 27,279 | -0.09(-0.35, 0.16) | 0.33(0.01, 0.65)* | 0.05(-0.19, 0.28) | 0.12(-0.01, 0.25) | -0.40(-0.48, -0.32)* |
| Short sleep | 15,739 | -0.49 (-1.14,0.15) | 0.37 (-0.20,0.94) | 0.27 (-0.04,0.58) | 0.21 (0.02, 0.40) | -0.36 (-0.47, -0.25)* |
| Long sleep | 11,540 | 1.06(0.10, 2.01)* | -0.14(-0.93, 0.65) | -0.38(-0.78, 0.01) | -0.09(-0.31, 0.13) | -0.44 (-0.55, -0.33)* |
| **Model 2: Major CVD mortality** | | | | | | |
| Meeting sleep guidelines | 28,653 | 1.92 (-0.22, 4.07) | -1.01(-2.62, 0.61) | -0.35(-1.08, 0.37) | -0.26(-0.70, 0.18) | -0.31(-0.54, -0.07)* |
| Not meeting sleep guidelines | 24,372 | -0.80 (-1.38, -0.21)* | 0.71(-0.01, 1.42) | 0.31(-0.21, 0.83) | 0.28(-0.02, 0.57) | -0.50(-0.67, -0.33)* |
| Short sleep | 14,184 | -1.48 (-2.86, -0.11)* | 1.14 (-0.09, 2.36) | 0.44 (-0.22,1.10) | 0.33 (-0.08, 0.74) | -0.42(-0.66, -0.19)* |
| Long sleep | 10,188 | -1.97(4.32, 0.39) | 1.62(-0.31, 3.56) | 0.43(-0.53, 1.39) | 0.46(-0.08, 1.01) | -0.55(-0.82, -0.29)* |
| **Model 3: PA-related cancer mortality** | | | | | | |
| Meeting sleep guidelines | 28,584 | 0.80 (-0.46, 2.07) | 0.22(-0.73, 1.18) | -0.59(-1.03, -0.15)* | -0.37(-0.64, -0.10)* | -0.07(-0.23, 0.09) |
| Not meeting sleep guidelines | 24,625 | 0.07 (-0.33, 0.47) | -0.19(-0.69, 0.29) | 0.20(-0.14, 0.55) | 0.12 (-0.08, 0.32) | -0.20(-0.33, -0.07)* |
| Short sleep | 14,415 | -0.12 (-1.14, 0.89) | -0.33 (-1.18,0.53) | 0.47 (-0.01,0.94) | 0.17 (-0.13,0.46) | -0.18 (-0.36, -0.003)* |
| Long sleep | 10,210 | 0.90(-0.54,2.34) | -0.38(-1.56, 0.81) | -0.26 (-0.84, 0.32) | -0.05(-0.38, 0.29) | -0.21(-0.40, -0.03)* |
| The results represent the association of one behaviour, relative to all others, on the outcome. Coefficients indicate change in log-hazard of mortality per 1 unit increase in the corresponding ILR coordinate; they indicate the presence of an association, but effect size is not directly interpretable due to the isometric log-ratio transformation. Value >0 indicates more time spent in the behaviour relative to others is associated with higher hazard; value <0 indicates that more time spent in the behaviour relative to others is associated with lower hazard.  * Indicates significant results at p<0.05 | | | | | | |

### eTable 4 Interaction test between the ILR coordinate for sleep versus all other behaviours and sleep characteristics on three mortality outcomes

| Interaction | P-value (All-cause) | P-value (CVD) | P-value (Cancer) |
| --- | --- | --- | --- |
| Sleep vs. all *Duration | <0.0001 | 0.1666 | 0.2497 |
| Sleep vs. all *Regularity | 0.9887 | 0.8324 | 0.7224 |

Models adjusted for age, sex, ethnicity, SRI, BMI, diet, smoking, alcohol consumption, tea and coffee consumption, education, employment shift, sleep problems, medications, parental history of CVD (all-cause and CVD morality models), parental history of cancer (all-cause and cancer morality models), and prevalent CVD and prevalent cancer (all-cause mortality model). P-values represent the significance of the interaction terms between the ILR coordinate for sleep relative to all other behaviours (Sleep vs. all) and sleep characteristics (duration and regularity) in their association with all-cause, CVD and cancer mortality.

### eTable 5 Baseline characteristics of participants stratified by sleep regularity index (SRI) group (n=58,149).

|  | Overall | Irregular (SRI ≤ 87.8) | Regular (SRI > 87.8) |
| --- | --- | --- | --- |
| n | 58,149 | 43,642 | 14,507 |
| Sleep (mean (SD)) | 7.6 (1.0) | 7.4 (1.0) | 8.0 (0.8) |
| Sedentary behaviour (mean (SD)) | 10.5 (1.3) | 10.6 (1.3) | 10.2 (1.2) |
| Standing (mean (SD)) | 2.9 (0.8) | 2.9 (0.8) | 2.9 (0.8) |
| LPA (mean (SD)) | 2.1 (1.0) | 2.1 (1.0) | 2.1 (1.0) |
| MVPA (mean (SD)) | 0.7 (0.4) | 0.7 (0.4) | 0.7 (0.4) |
| Age (mean (SD)) | 62.2 (7.8) | 62.0 (7.8) | 62.7 (7.8) |
| Sex = Male (%) | 25,519 (43.9) | 18,656 (42.7) | 6,863 (47.3) |
| Ethnicity = White (%) | 54,598 (93.9) | 40,730 (93.3) | 13,868 (95.6) |
| BMI (mean (SD)) | 26.6 (4.5) | 26.9 (4.6) | 25.9 (4.0) |
| Fruit and vegetable intake^1^ (mean (SD)) | 8.0 (4.4) | 8.0 (4.5) | 8.0 (4.3) |
| Smoking status (%) |  | | |
| Current | 3,600 (6.2) | 2,977 (6.8) | 623 (4.3) |
| Never | 33,289 (57.2) | 24,479 (56.1) | 8,810 (60.7) |
| Previous | 21,260 (36.6) | 16,186 (37.1) | 5,074 (35.0) |
| Alcohol consumption^2^ (mean (SD)) | 13.7 (15.2) | 13.9 (15.5) | 13.3 (14.2) |
| Coffee and tea intake, cups per day (mean (SD)) | 5.3 (2.8) | 5.3 (2.8) | 5.3 (2.6) |
| Education^3^ (%) |  | | |
| A/AS level | 7,600 (15.1) | 5,625 (15.0) | 1,975 (15.5) |
| College | 25,272 (50.3) | 18,595 (49.5) | 6,677 (52.5) |
| CSE | 2,260 (4.5) | 1,814 (4.8) | 446 (3.5) |
| NVQ/HND/HNC | 3,245 (6.5) | 2,493 (6.6) | 752 (5.9) |
| O level | 11,907 (23.7) | 9,044 (24.1) | 2,863 (22.5) |
| Employment shift^4^ (%) |  | | |
| Employed in day shift work | 2,251 (3.9) | 1,836 (4.2) | 415 (2.9) |
| Employed in night shift work | 1,945 (3.3) | 1,674 (3.8) | 271 (1.9) |
| Employed not in shift work | 29,378 (50.5) | 22,110 (50.7) | 7,268 (50.1) |
| Retired/not in workforce | 24,575 (42.3) | 18,022 (41.3) | 6,553 (45.2) |
| Healthy sleep score^5^ (mean (SD)) | 3.7 (1.0) | 3.7 (1.0) | 3.8 (1.0) |
| Medication use^6^ = Yes (%) | 13,640 (23.5) | 10,458 (24.0) | 3,182 (21.9) |
| Family history of CVD = Yes (%) | 32,326 (55.6) | 24,208 (55.5) | 8,118 (56.0) |
| Family history of cancer = Yes (%) | 18,049 (31.0) | 13,474 (30.9) | 4,575 (31.5) |

The columns breakdown corresponds to irregular and regular sleepers: irregular sleepers, SRI≤87.8; regular sleepers, SRI>87.8. Values represent mean (SD) unless specified otherwise.

^1^Fruits and vegetable consumption is servings per day.

^2^Alcohol consumption: above guidelines are >14 units per week, where 1 unit = 8 g of ethanol.

^3^A/AS level.; CSE,; Higher National Diploma,; IQR, interquartile range; National Vocational Qualification,; O level.

^4^Shift work was defined as a work schedule that falls outside of the normal daytime working hours of 9am-5pm. Night shifts are identified when the corresponding work schedule involves working through the normal sleeping hours, for instance working through the hours from 12am to 6am.

^5^Composited sleep score of self-reported chronotype, sleep duration, insomnia, snoring and daytime sleepiness, scoring from 0 (poor sleep) to 5 (healthy sleep).

^6^Medication use for cholesterol, diabetes or blood pressure.

eFigure 1: Study participant flow diagram

Participants with physical intensity accelerometry data

(n=72,581)

Participants with insufficient days of valid accelerometry wear time (<5 weekdays and <1 weekend day) (n=1,540)

Participants with sufficient valid accelerometry data (n= 71,041)

Participants with missing covariate data (n=12,779):

age (n = 65), sex (n = 65), ethnicity (n = 255), BMI (n = 925), diet (n = 65), smoking (n = 193), alcohol intake (n = 193), tea/coffee intake (n = 131), education (n = 217), shift work status (n = 65), sleep score (n = 11554), medication use (n = 545), parental history of CVD or cancer (n = 65)

Participants with complete data (n= 58,262)

Participants with a mortality event within 12 months of follow-up (n=113)

Sample for total mortality

(n= 58,149; 2209 events)

Participants with prevalent CVD (n=5,124)

Participants with prevalent cancer (n=4,940)

Sample for cancer mortality

(n= 53,209; 997 events)

Sample for CVD mortality

(n= 53,025; 435 events)

### eFigure 2: Sample mean composition among 58,149 participants.

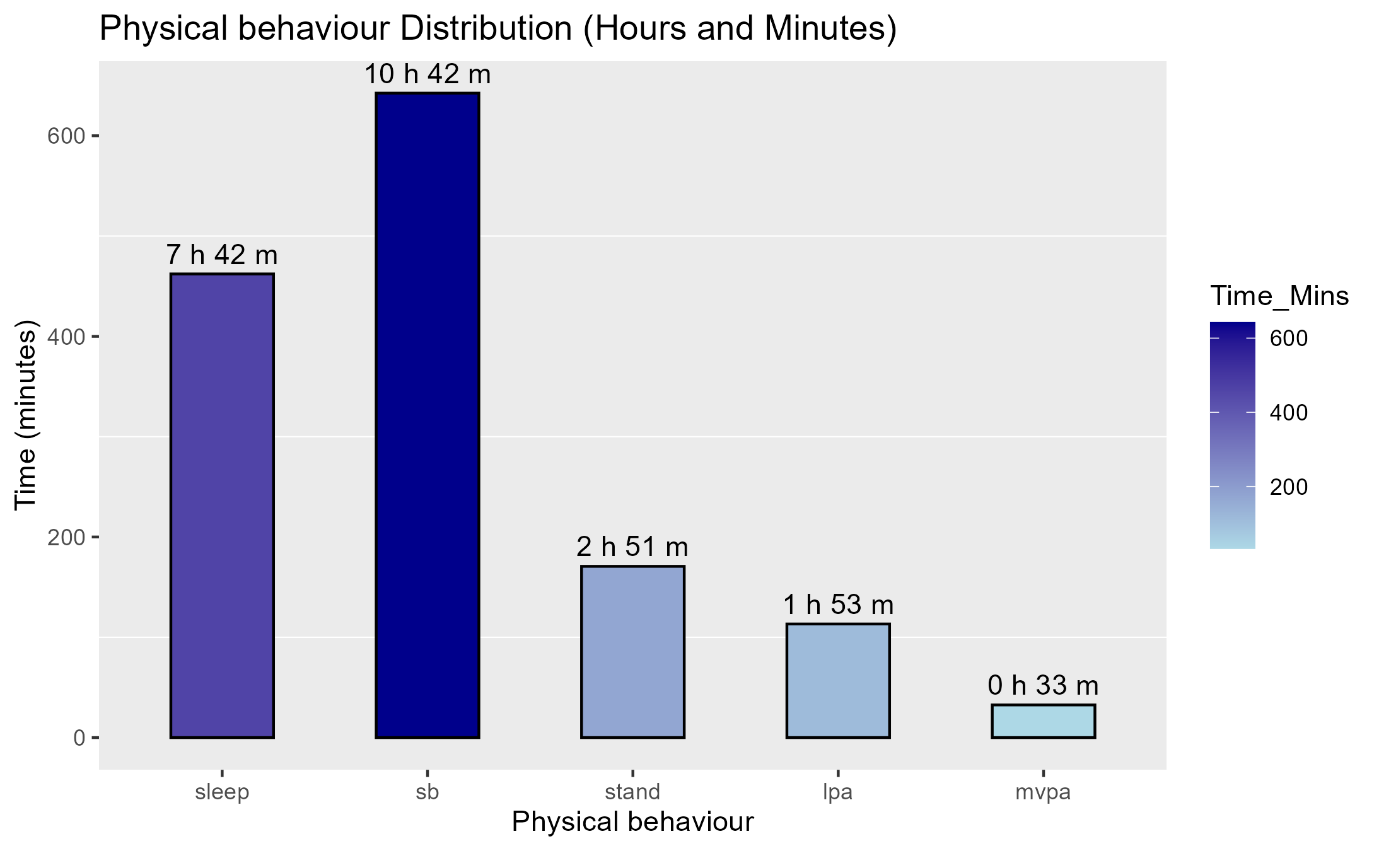
Compositional mean representing the hypothetical average daily time distribution of an individual’s physical behaviours.

### eFigure 3: Relative difference in time (expressed as log-ratio difference) spent in each behaviour in comparison to the overall sample mean composition (by mortality status)

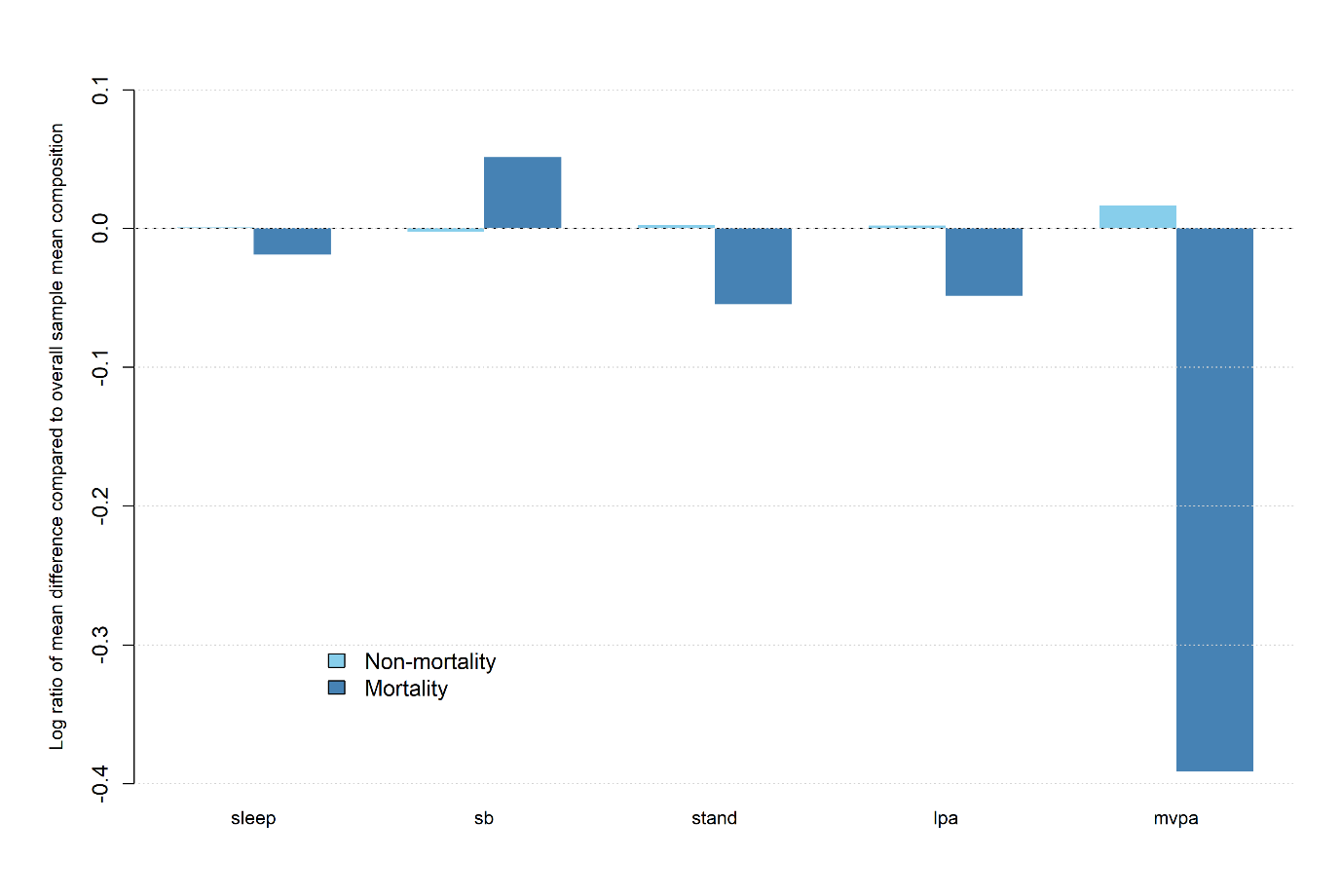

### eFigure 4: Associations of time reallocation between sleep and other behaviours with mortality outcomes

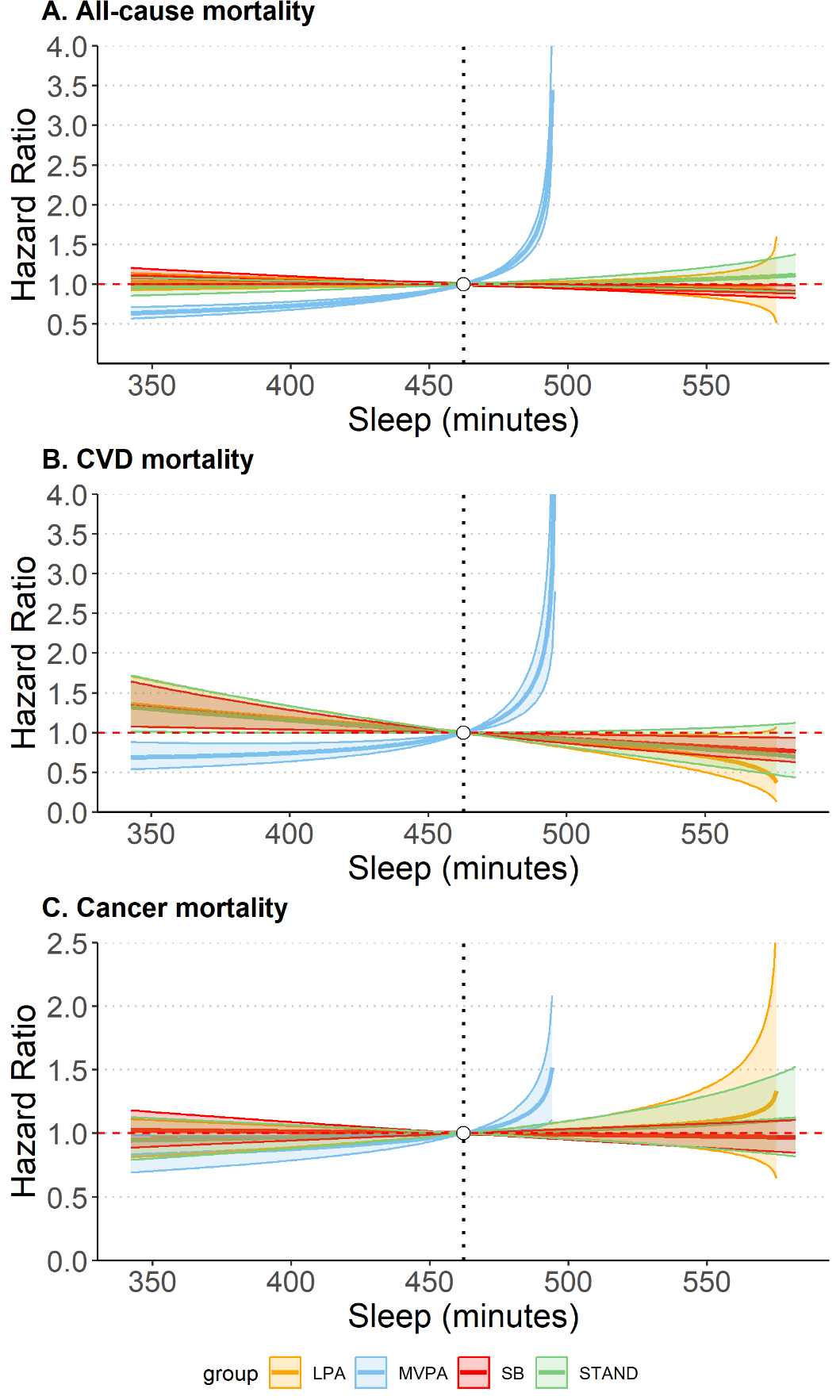

Substitution models for sleep for A) all-cause mortality (n=58,149, 2,209 events); B) CVD mortality (n=53,025, 683 events); C) cancer mortality (n=53,209, 997 events). Models adjusted for all covariates. Models adjusted for all covariates.

### eFigure 5: Associations of time reallocation between physical behaviours with all-cause mortality

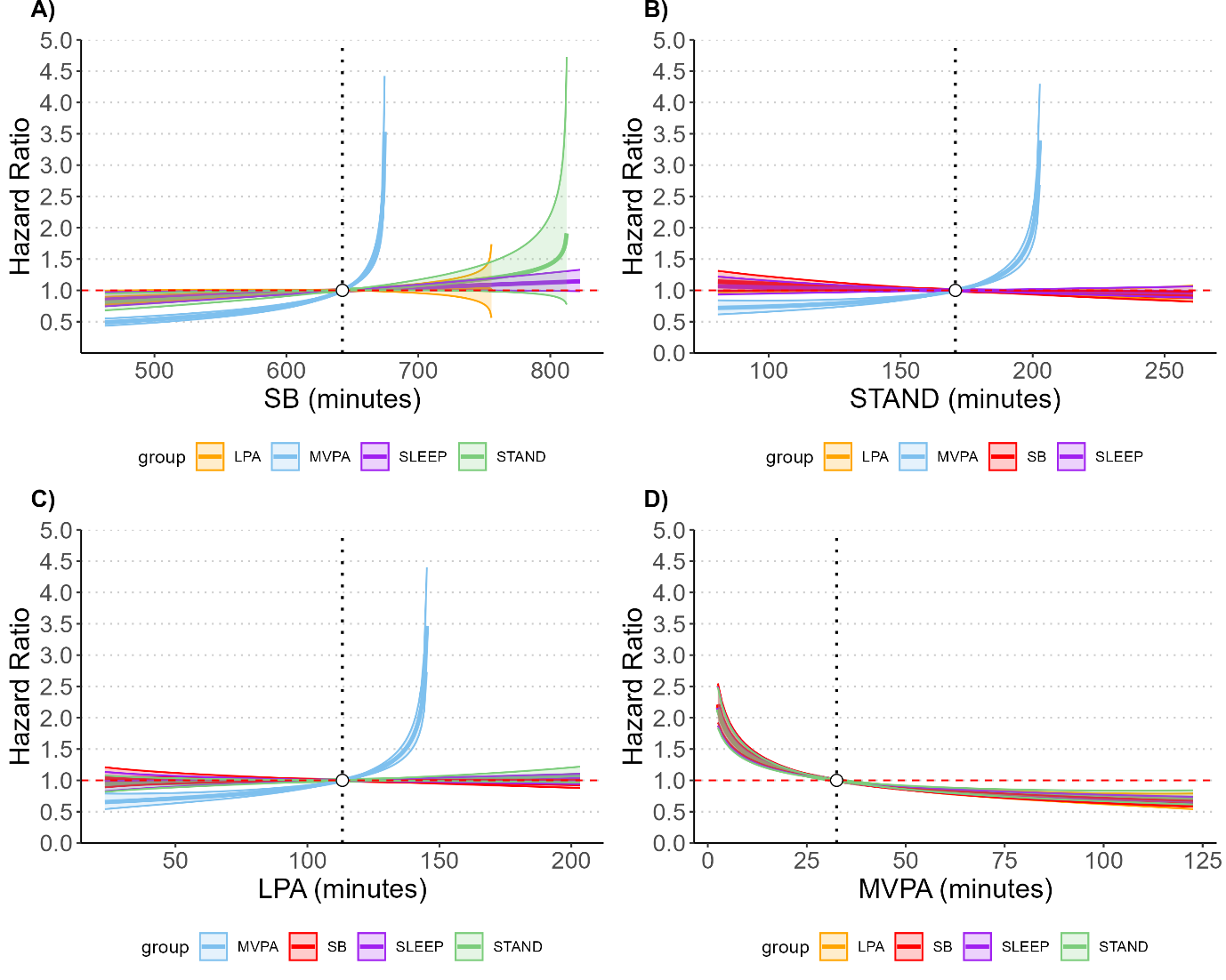

Substitution models (n=58,149, 2,209 events) for all-cause mortality for A) sedentary behaviour (SB); B) standing; C) light intensity physical activity (LPA); D) moderate to vigorous physical activity (MVPA). Models adjusted for all covariates.

### eFigure 6: Associations of time reallocation between physical behaviours with CVD mortality

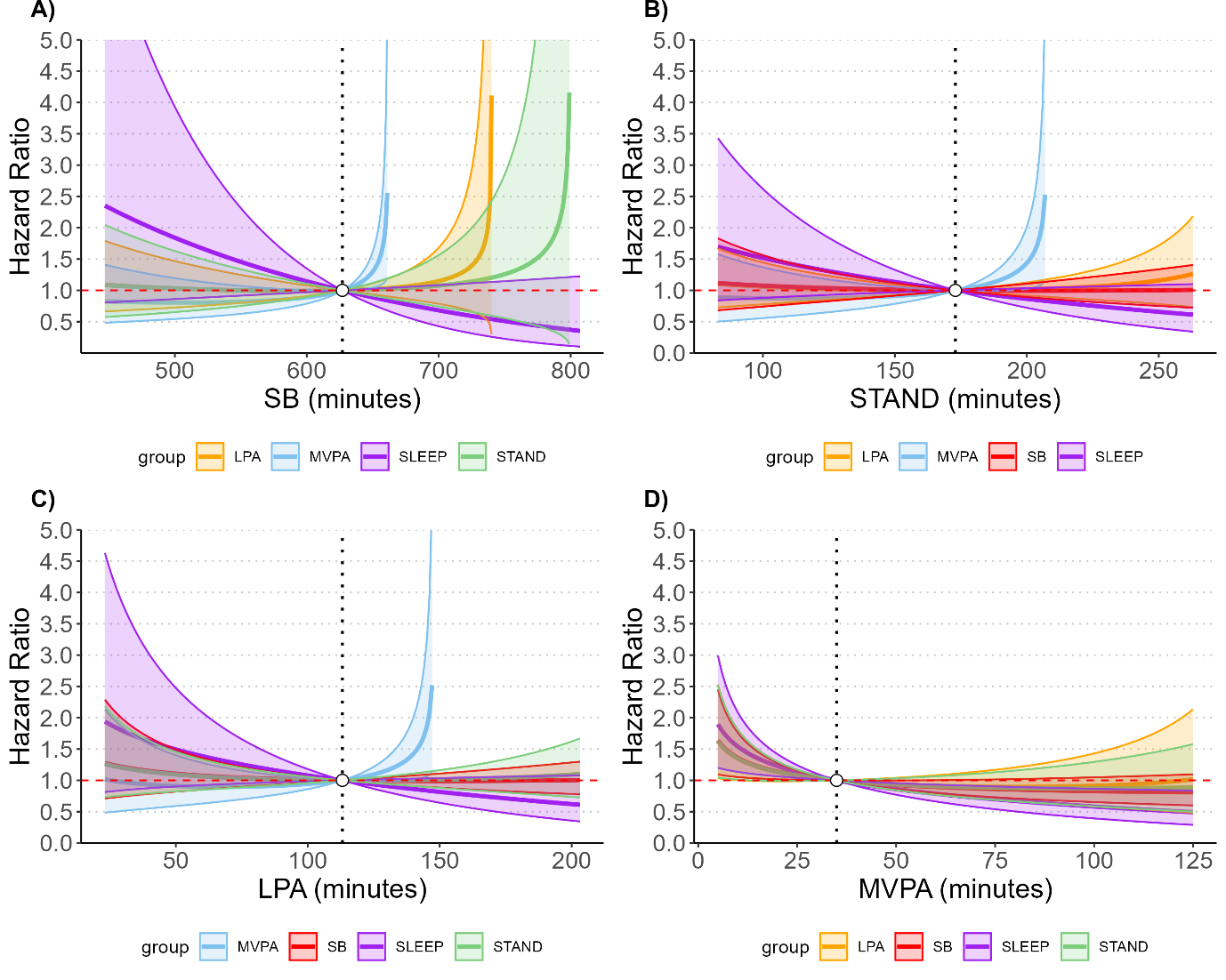

Substitution models (n=53,025, 683 events) for CVD mortality for A) sedentary behaviour (SB); B) standing; C) light intensity physical activity (LPA); D) moderate to vigorous physical activity (MVPA). Models adjusted for all covariates.

### eFigure 7: Associations of time reallocation between physical behaviours with Cancer mortality

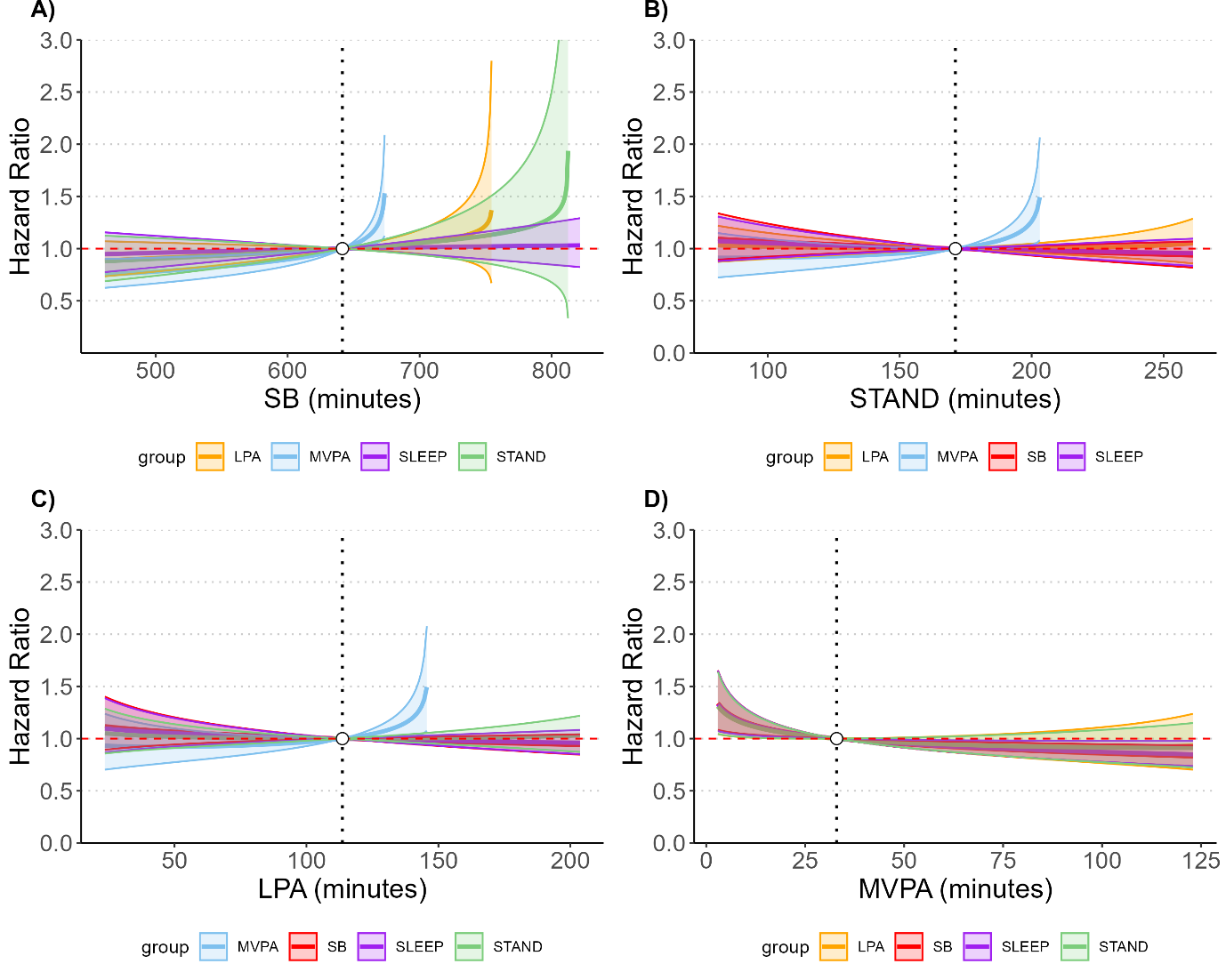

Substitution models (n=53,209, 997 events) for cancer mortality for A) sedentary behaviour (SB); B) standing; C) light intensity physical activity (LPA); D) moderate to vigorous physical activity (MVPA). Models adjusted for all covariates.

### eFigure 8: Associations of time reallocation between physical behaviours with all-cause mortality stratified by sleep duration

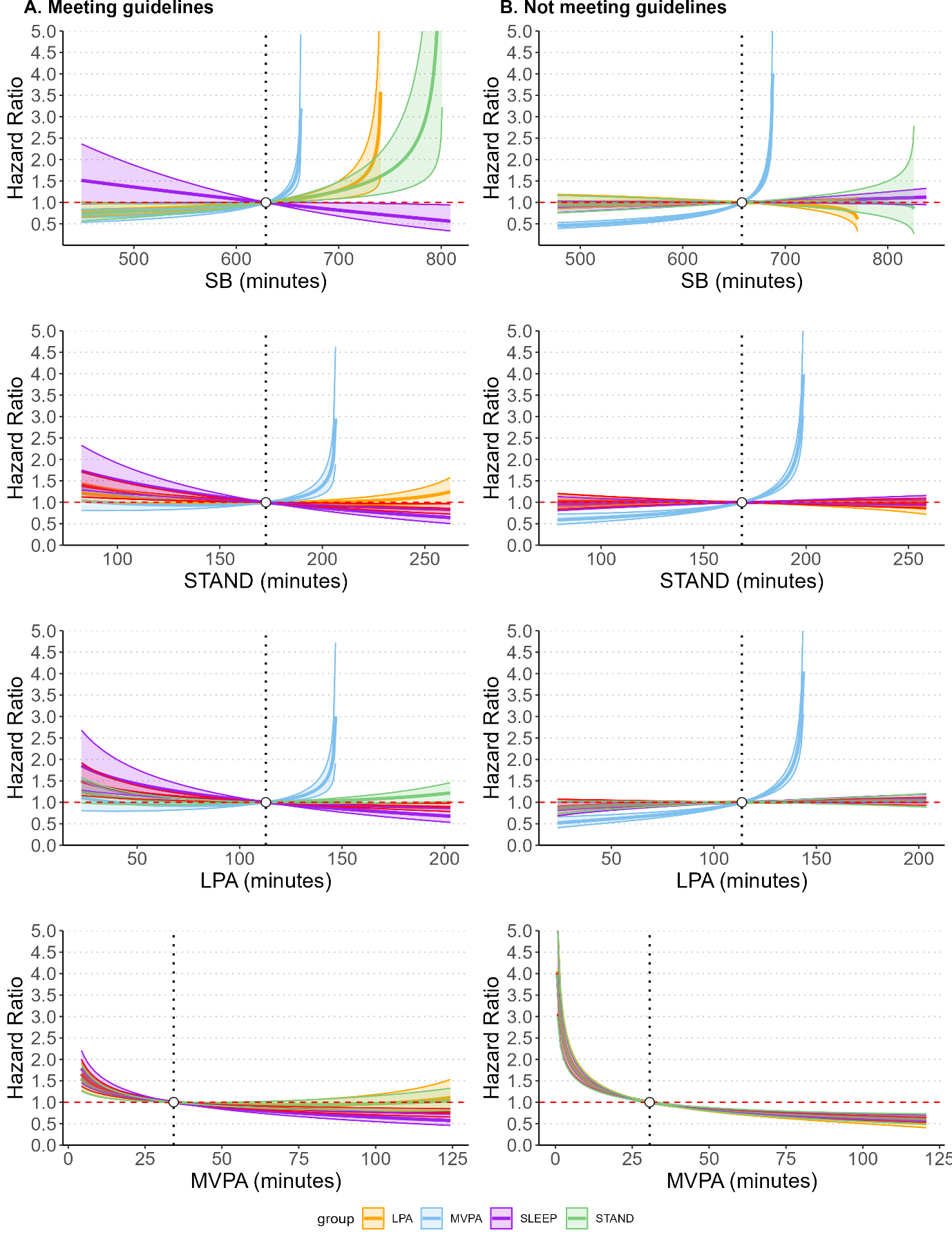

Substitution models for all-cause mortality for participants A. meeting sleep duration guidelines (n=30,870; 901 events) with reference compositional mean of 10.5h SB, 2.9h standing, 1.9h LPA, and 0.6h MVPA; B. not meeting guidelines (n=27,279, 1,308 events) with reference compositional mean of 11.0h SB, 2.8h standing, 1.9h LPA, 0.5h MVPA. Models adjusted for all covariates.

### eFigure 9: Associations of time reallocation between physical behaviours with CVD mortality stratified by sleep duration

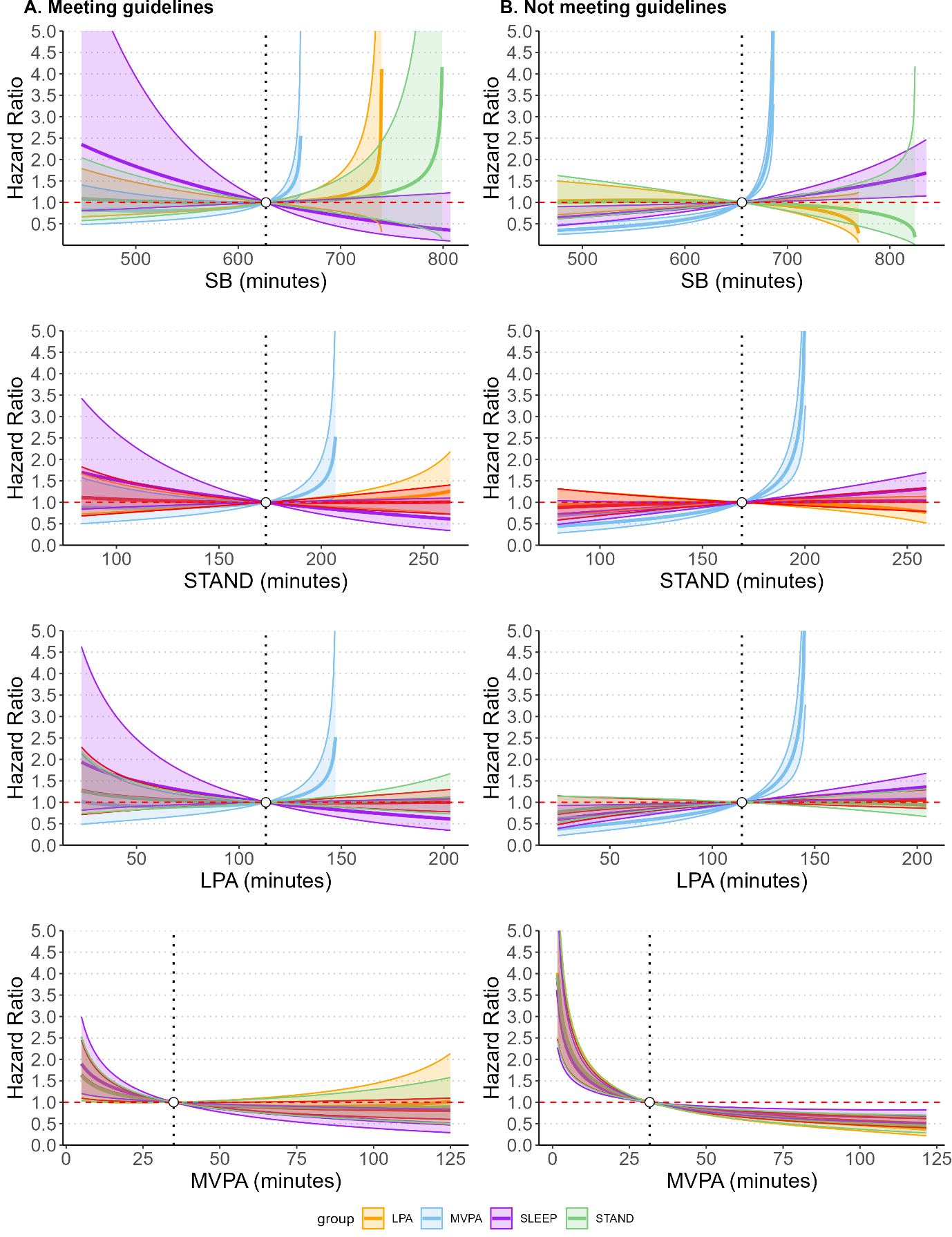

Substitution models for all-cause mortality for participants A. meeting sleep duration guidelines (n=28,653; 169 events) with reference compositional mean of 10.4h SB, 2.9h standing, 1.9h LPA, and 0.6h MVPA; B. not meeting guidelines (n=24,372, 266 events) with reference compositional mean of 10.9h SB, 2.8h standing, 1.9h LPA, 0.5h MVPA. Models adjusted for all covariates.

### eFigure 10: Associations of time reallocation between physical behaviours with cancer mortality stratified by sleep duration

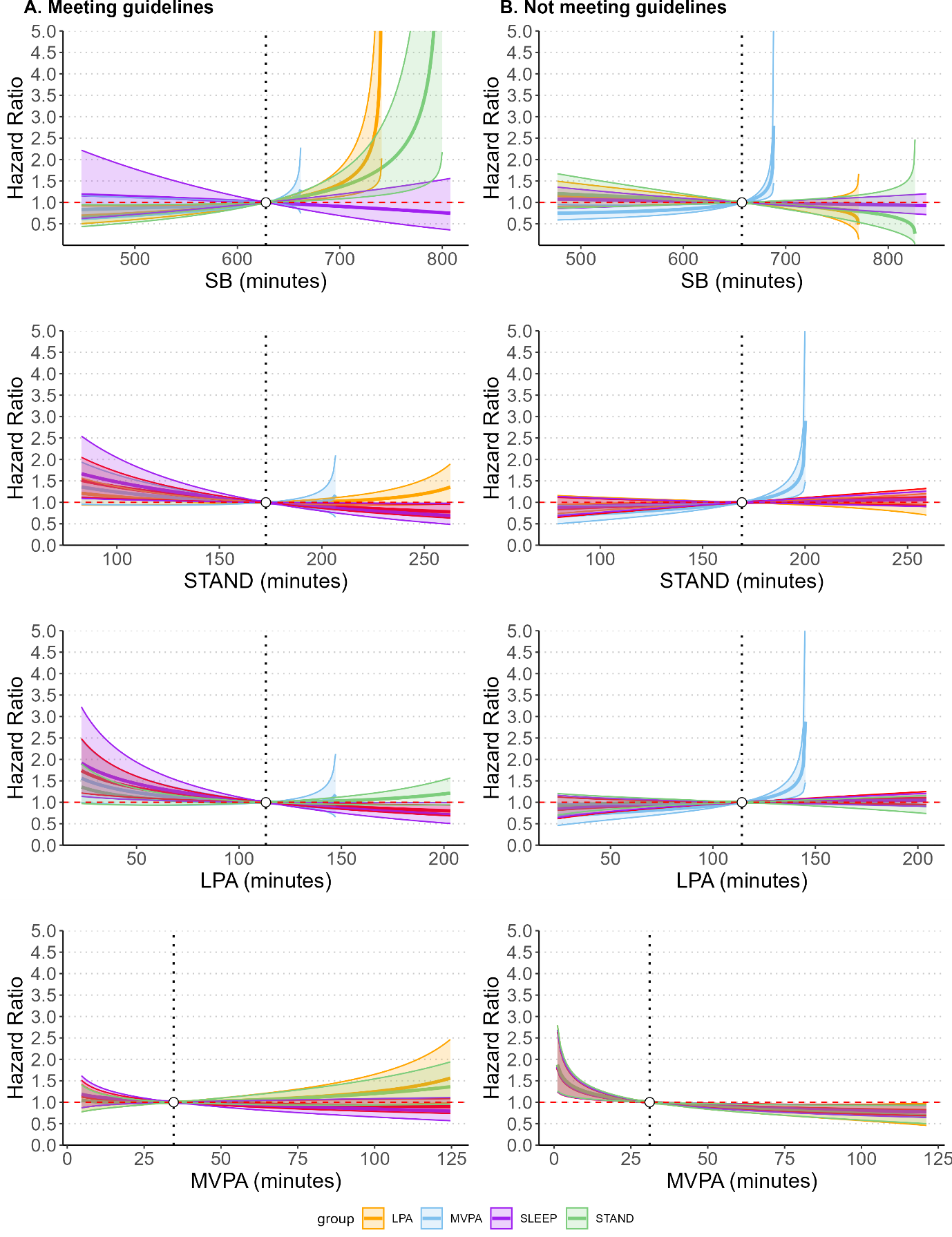

Substitution models for all-cause mortality for participants A. meeting sleep duration guidelines (n=28,584; 435 events) with reference compositional mean of 10.5h SB, 2.9h standing, 1.9h LPA, and 0.6h MVPA; B. not meeting guidelines (n=24,625, 562 events) with reference compositional mean of 11.0h SB, 2.8h standing, 1.9h LPA, 0.5h MVPA. Models adjusted for all covariates.

### eFigure 11: Associations of time reallocation between physical behaviours with CVD and cancer mortality stratified by sleep regularity

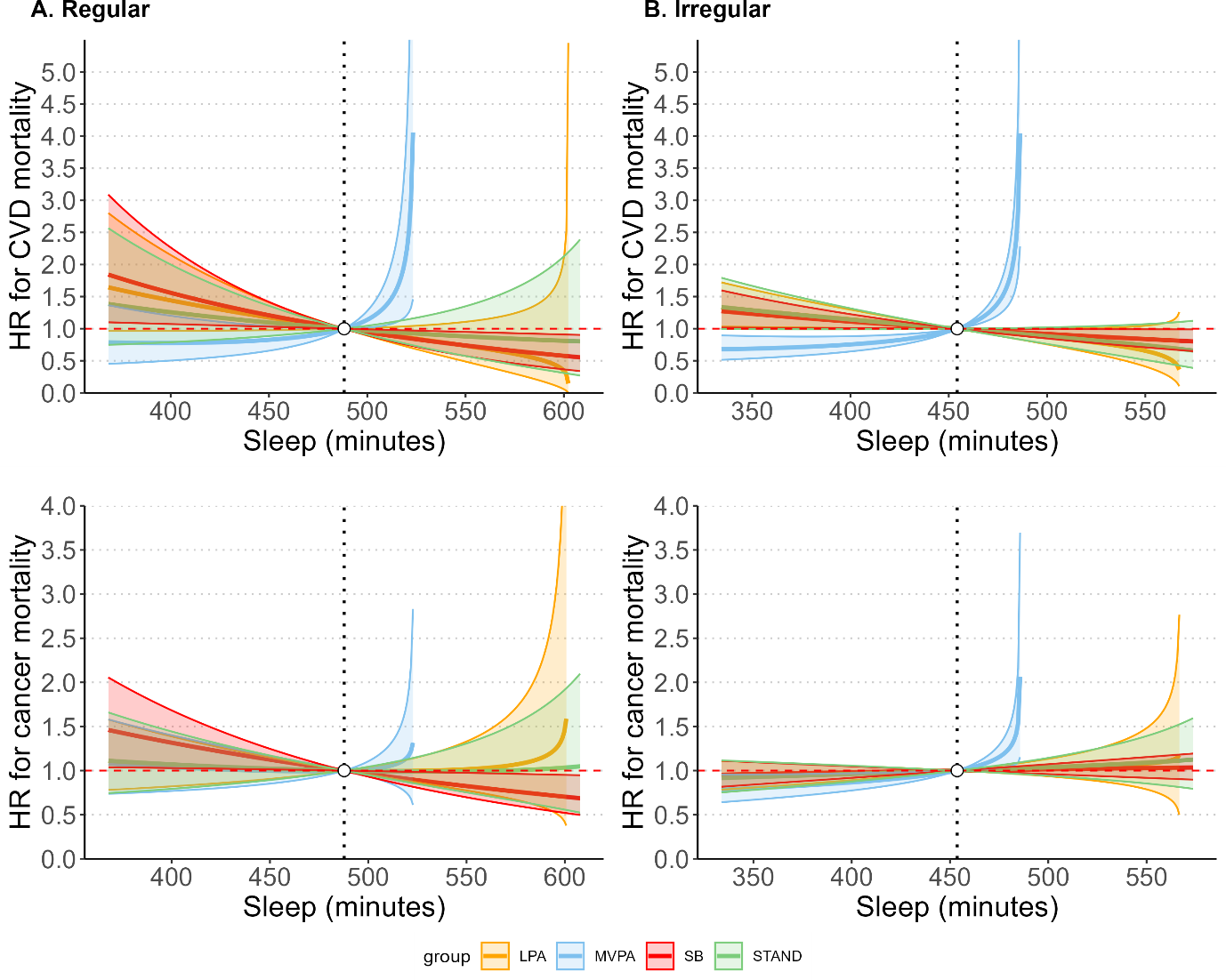

Estimated hazard ratios with 95%CIs in CVD and cancer mortality associated with the reallocation of time between sleep and other behaviours across sleep regularity groups: A. regular sleep (n=14,507; 517 events); B. irregular sleep (n=43,642; 1,692 events). Reference (dashed vertical line) is the sample compositional mean sleep duration: A. 488min (8.13h), B. 454min (7.56h). To the left of the reference shows replacing sleep with other behaviours, while to the right shows replacing other behaviours with sleep. Models adjusted for age, sex, ethnicity, BMI, diet, smoking, alcohol consumption, tea and coffee consumption, education, employment shift, sleep problems, medications, parental history of CVD (CVD model), parental history of cancer (cancer model).

### eFigure 12: Associations of time reallocation between physical behaviours with all-cause mortality stratified by sleep regularity

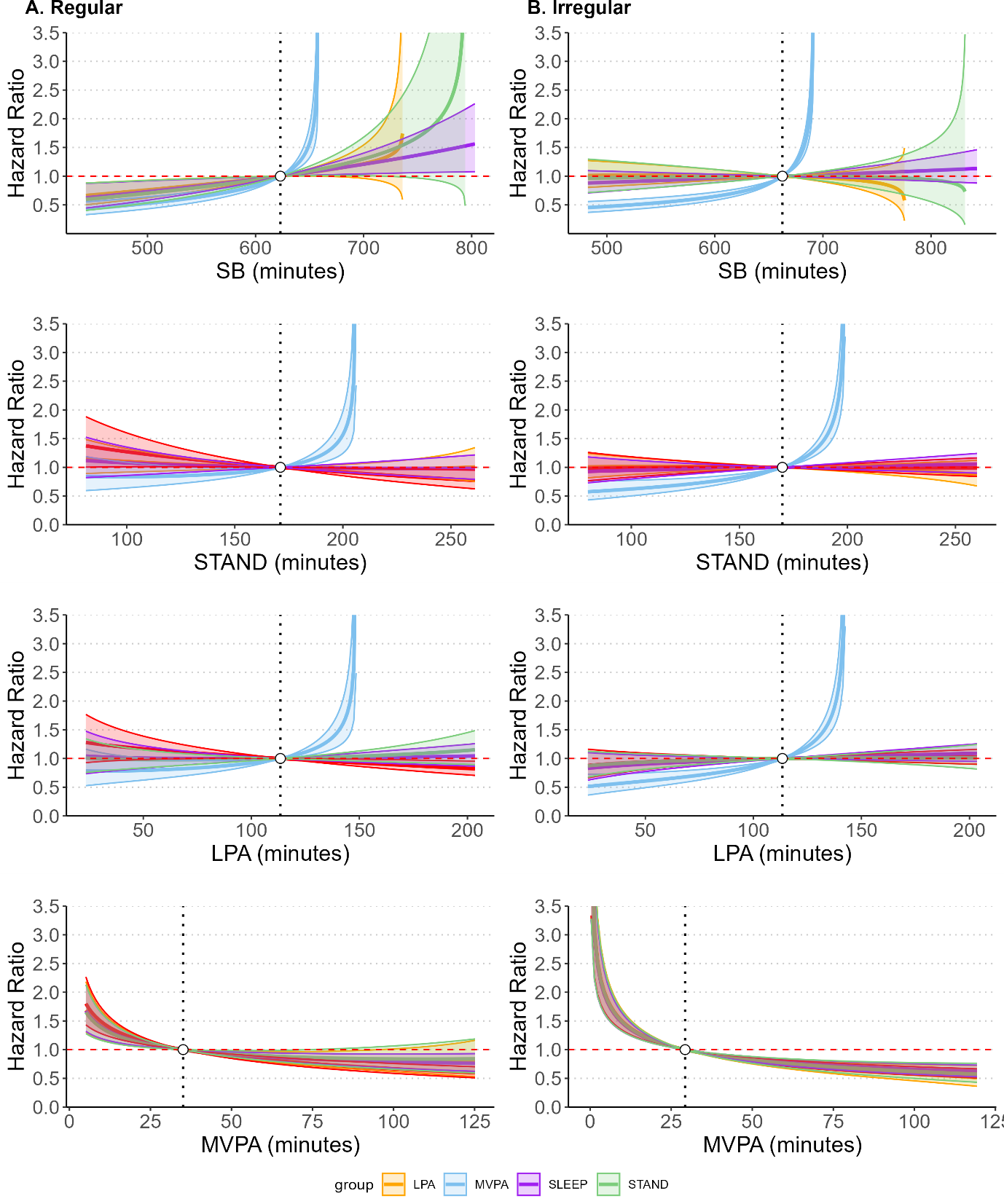

Substitution models for all-cause mortality for participants with A. regular sleep (n=14,507; 517 events) with reference compositional mean of 10.4h SB, 2.9h standing, 1.9h LPA, and 0.6h MVPA; B. and B. irregular sleep (n=43,642; 1,692 events) with reference compositional mean of 11.0h SB, 2.8h standing, 1.9h LPA, 0.5h MVPA. Models adjusted for all covariates.

### eFigure 13: Associations of time reallocation between sleep and other movement behaviours with all-cause mortality stratified by sleep duration excluding 287 events during first two years of follow-up

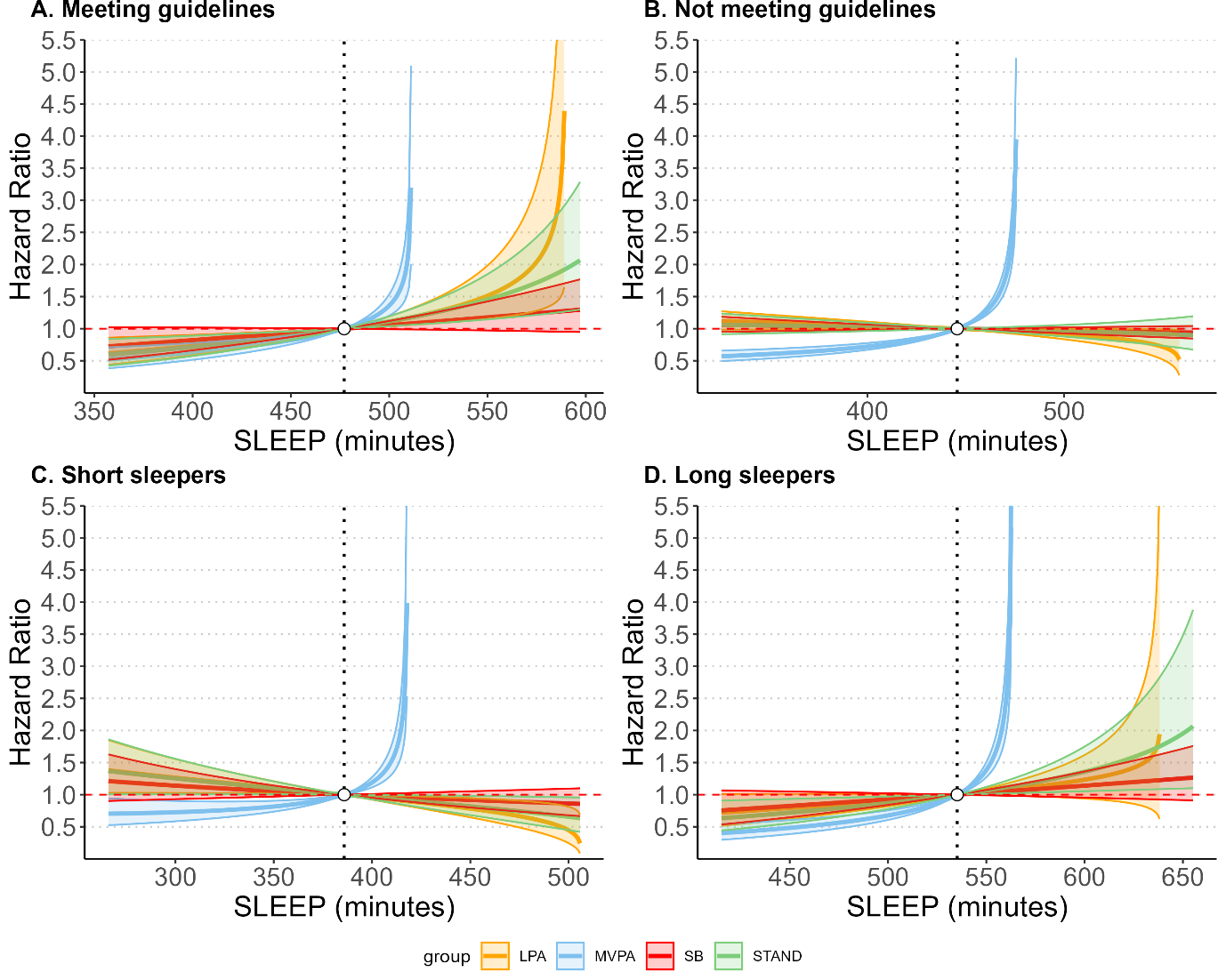

Estimated hazard ratios with 95%CIs in all-cause mortality associated with the reallocation of time between sleep and other behaviours across four sleep duration groups: A. meeting sleep duration guidelines (n=30,797; 828 events); B. not meeting guidelines (n=27,178; 1,207 events); C. short sleep (below guidelines) (n=15,684; 656 events); D. long sleep (above guidelines) (n=11,494; 551 events).

### eFigure 14: Associations of time reallocation between sleep and other movement behaviours with CVD mortality stratified by sleep duration excluding 287 events during first two years of follow-up

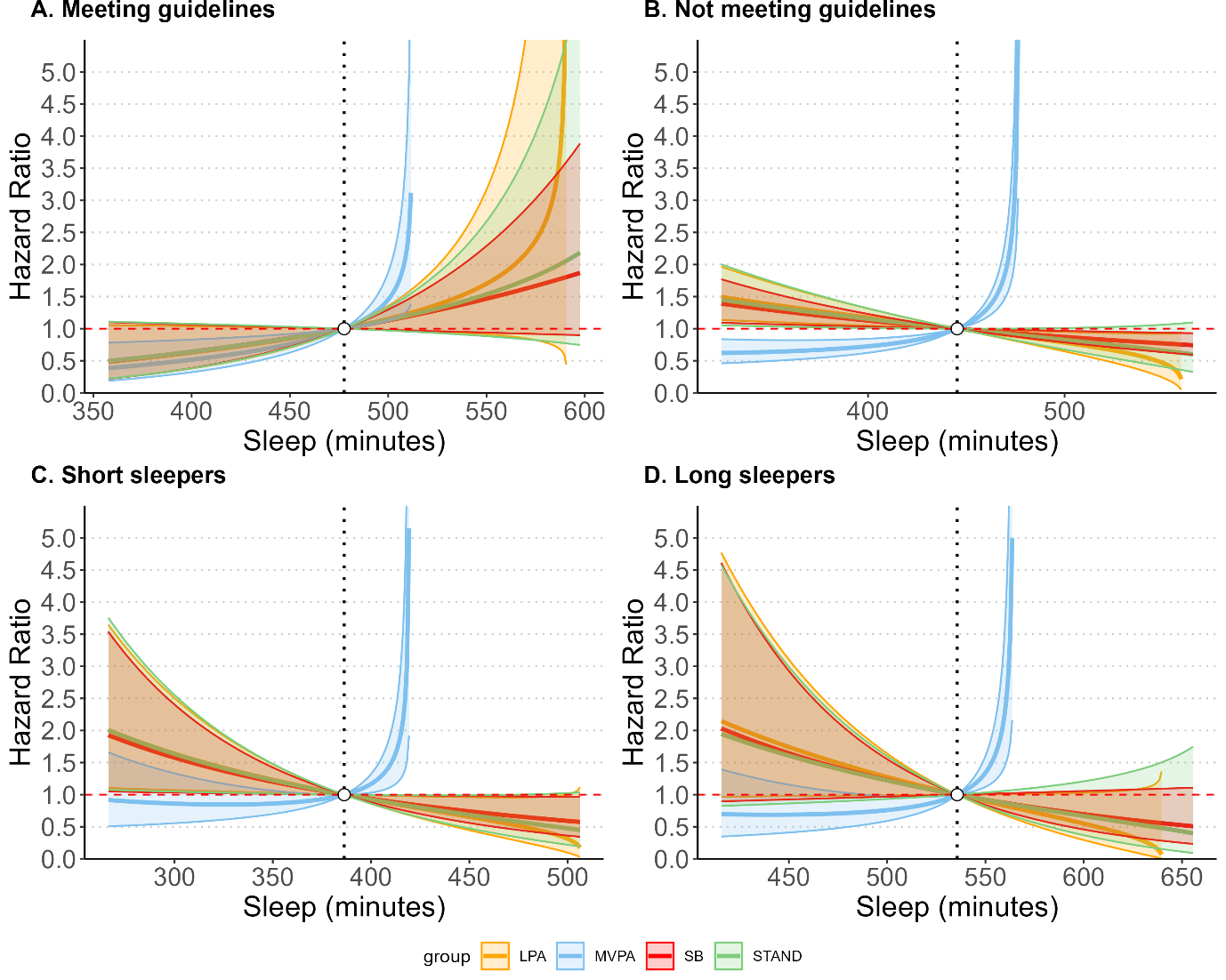

Estimated hazard ratios with 95%CIs in CVD mortality associated with the reallocation of time between sleep and other behaviours across four sleep duration groups: A. meeting sleep duration guidelines (7–9 hours/day for ages 18–64 and 7–8 hours/day for ages ≥65) (n=28,600; 162 events); B. not meeting guidelines (n=24,372; 266 events); C. short sleep (below guidelines) (n=14,184; 156 events); D. long sleep (above guidelines) (n=10,188; 110 events).

### eFigure 15: Associations of time reallocation between sleep and other movement behaviours with cancer mortality stratified by sleep duration excluding 287 events during first two years of follow-up

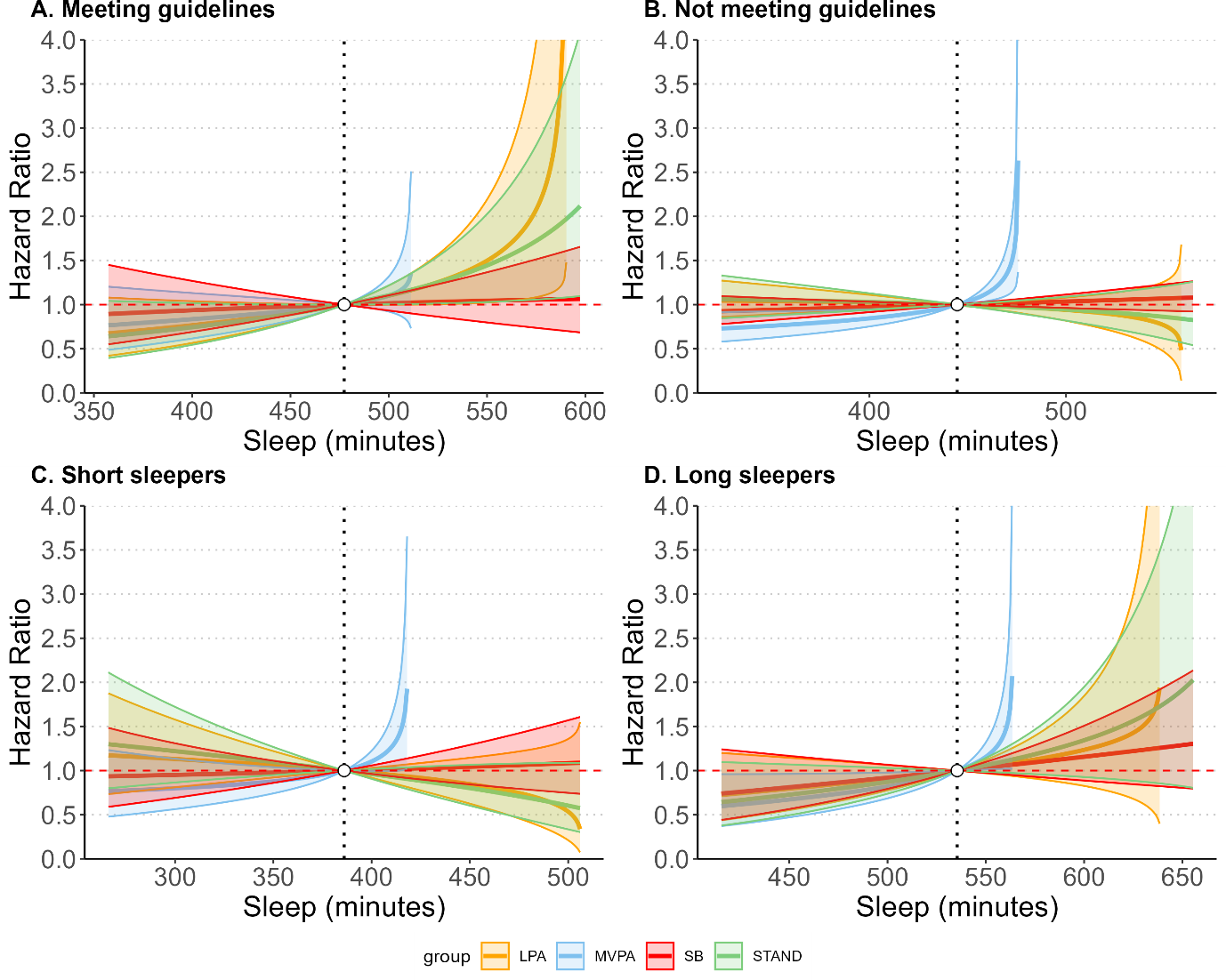

Estimated hazard ratios with 95%CIs in cancer mortality associated with the reallocation of time between sleep and other behaviours across four sleep duration groups: A. meeting sleep duration guidelines (7–9 hours/day for ages 18–64 and 7–8 hours/day for ages ≥65) (n=28,527; 402 events); B. not meeting guidelines (n=24,555; 520 events); C. short sleep (below guidelines) (n=14,373; 273 events); D. long sleep (above guidelines) (n=10,182; 247 events).

### eFigure 16: Associations of time reallocation between sleep and other movement behaviours with all-cause mortality stratified by sleep regularity excluding 287 events during first two years of follow-up

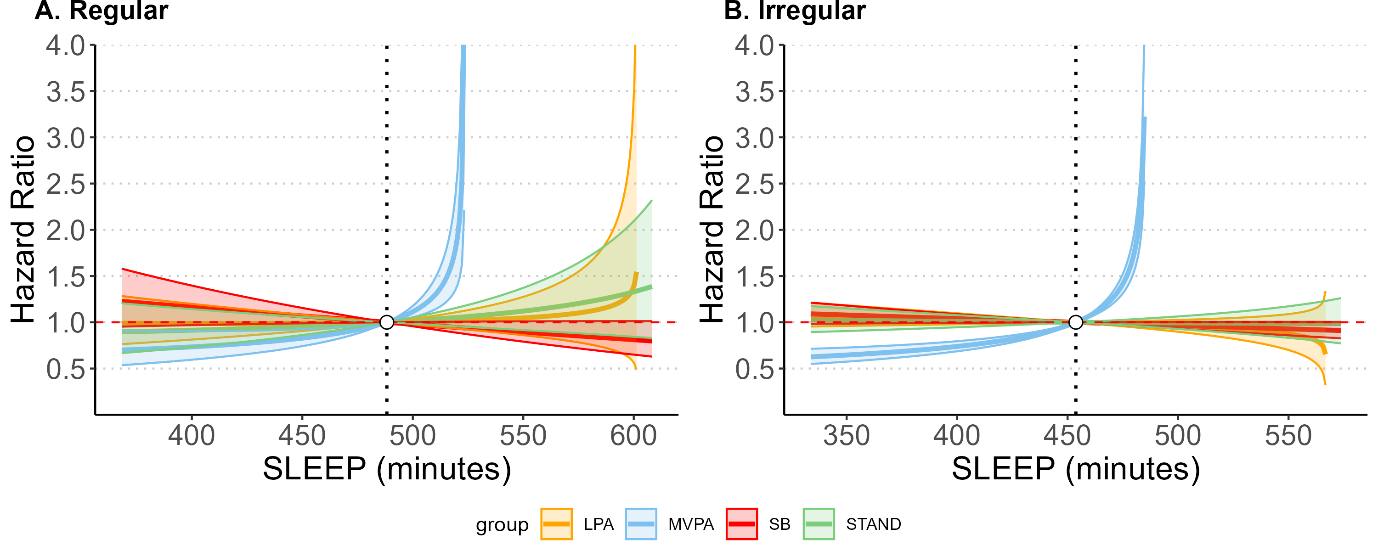

Estimated hazard ratios with 95%CIs in all-cause mortality associated with the reallocation of time between sleep and other behaviours across sleep regularity groups: A. regular sleep (n=14,454; 464 events); B. irregular sleep (n=43,521; 1,571 events).

### eFigure 17: Associations of time reallocation between sleep and other movement behaviours with all-cause mortality stratified by sleep duration excluding 1,120 participants with poor sleep

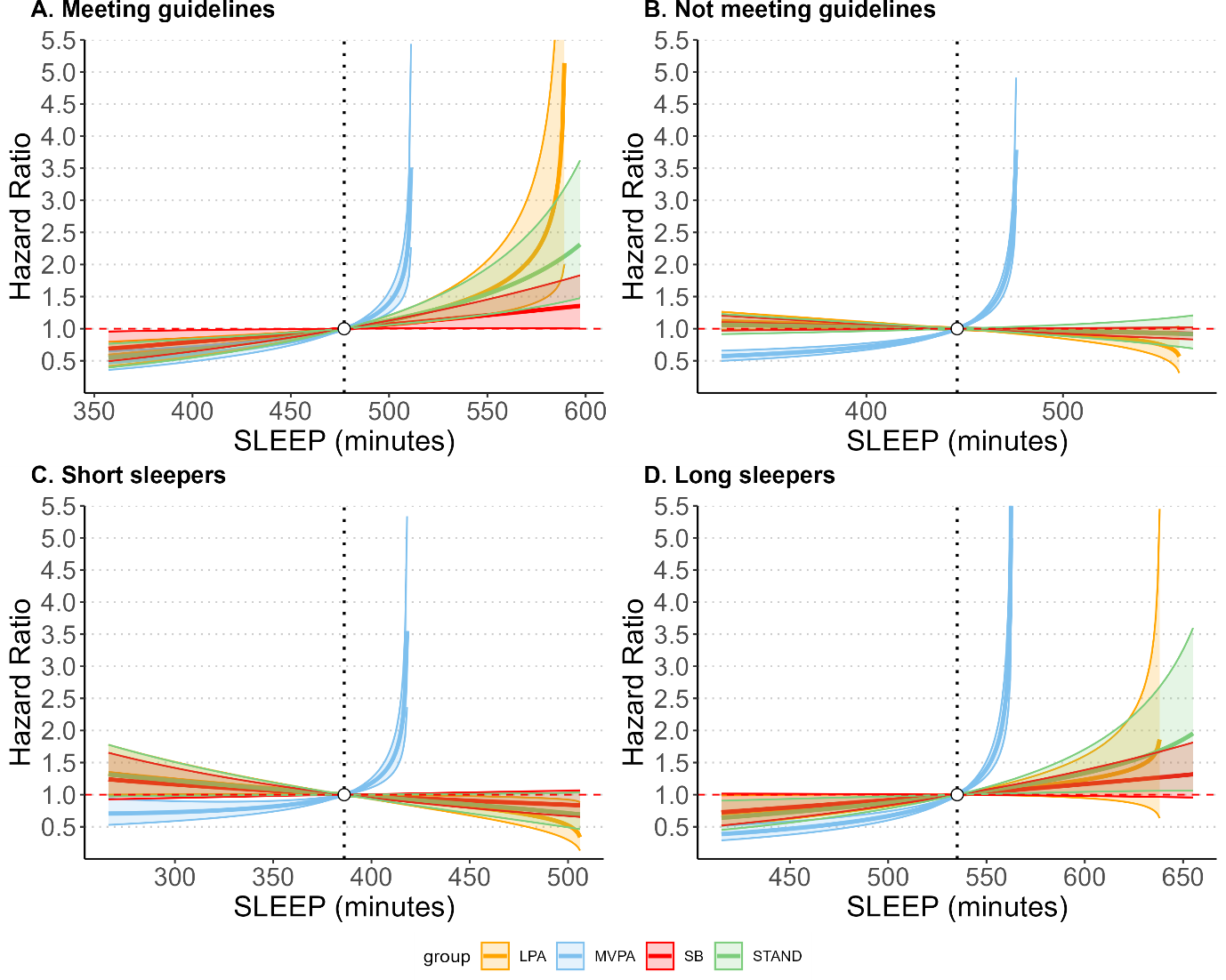

Estimated hazard ratios with 95%CIs in all-cause mortality associated with the reallocation of time between sleep and other behaviours across four sleep duration groups: A. meeting sleep duration guidelines (n=30,410; 883 events); B. not meeting guidelines (n=26,619; 1274 events); C. short sleep (below guidelines) (n=15,281; 691 events); D. long sleep (above guidelines) (n=11,338; 583 events).

### eFigure 18: Associations of time reallocation between sleep and other movement behaviours with CVD mortality stratified by sleep duration excluding 1,120 participants with poor sleep

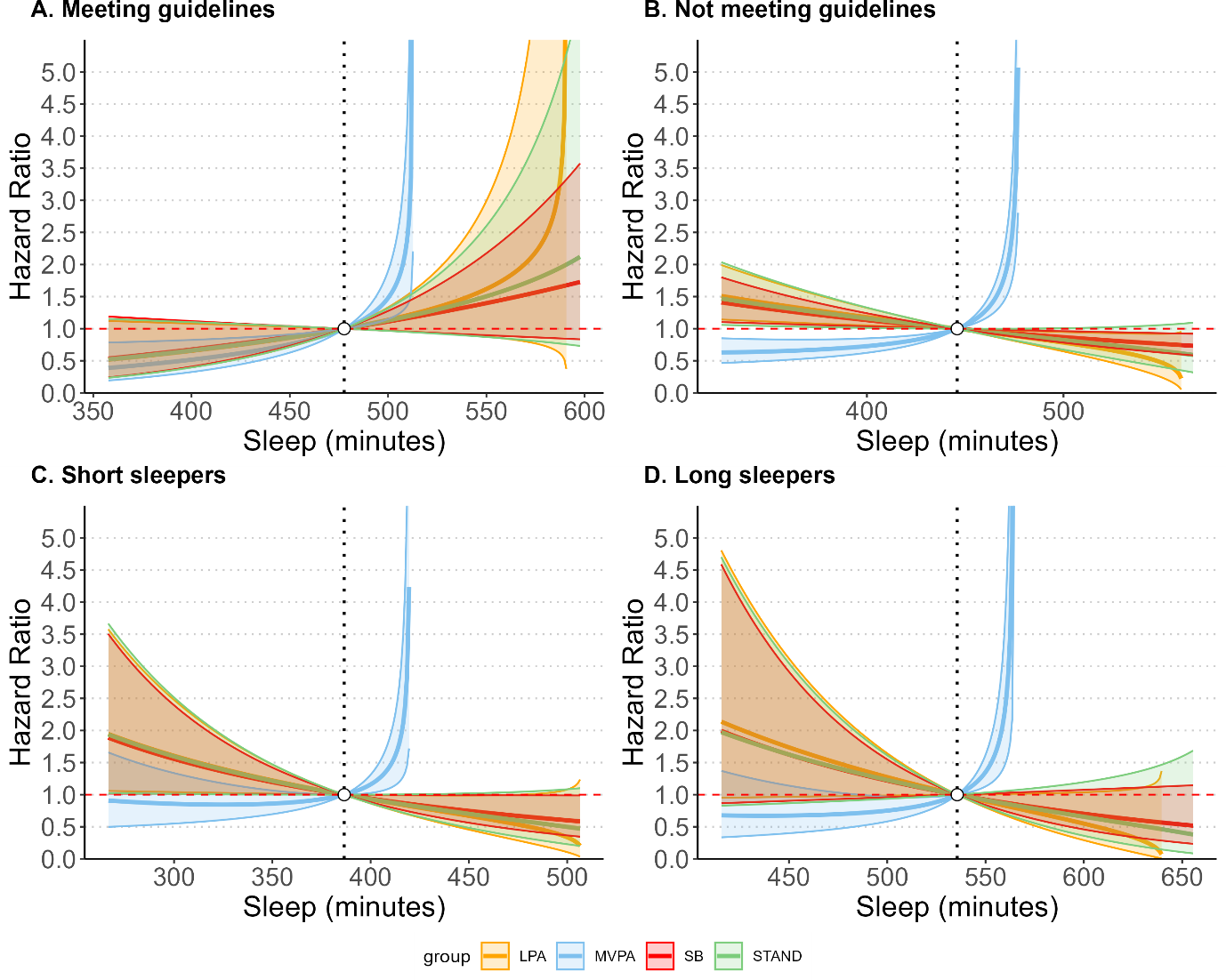

Estimated hazard ratios with 95%CIs in CVD mortality associated with the reallocation of time between sleep and other behaviours across four sleep duration groups: A. meeting sleep duration guidelines (n=28,250; 165 events); B. not meeting guidelines (n=23,824; 259 events); C. short sleep (below guidelines) (n=13,799; 152 events); D. long sleep (above guidelines) (n=10,025; 107 events).

### eFigure 19: Associations of time reallocation between sleep and other movement behaviours with cancer mortality stratified by sleep duration excluding 1,120 participants with poor sleep

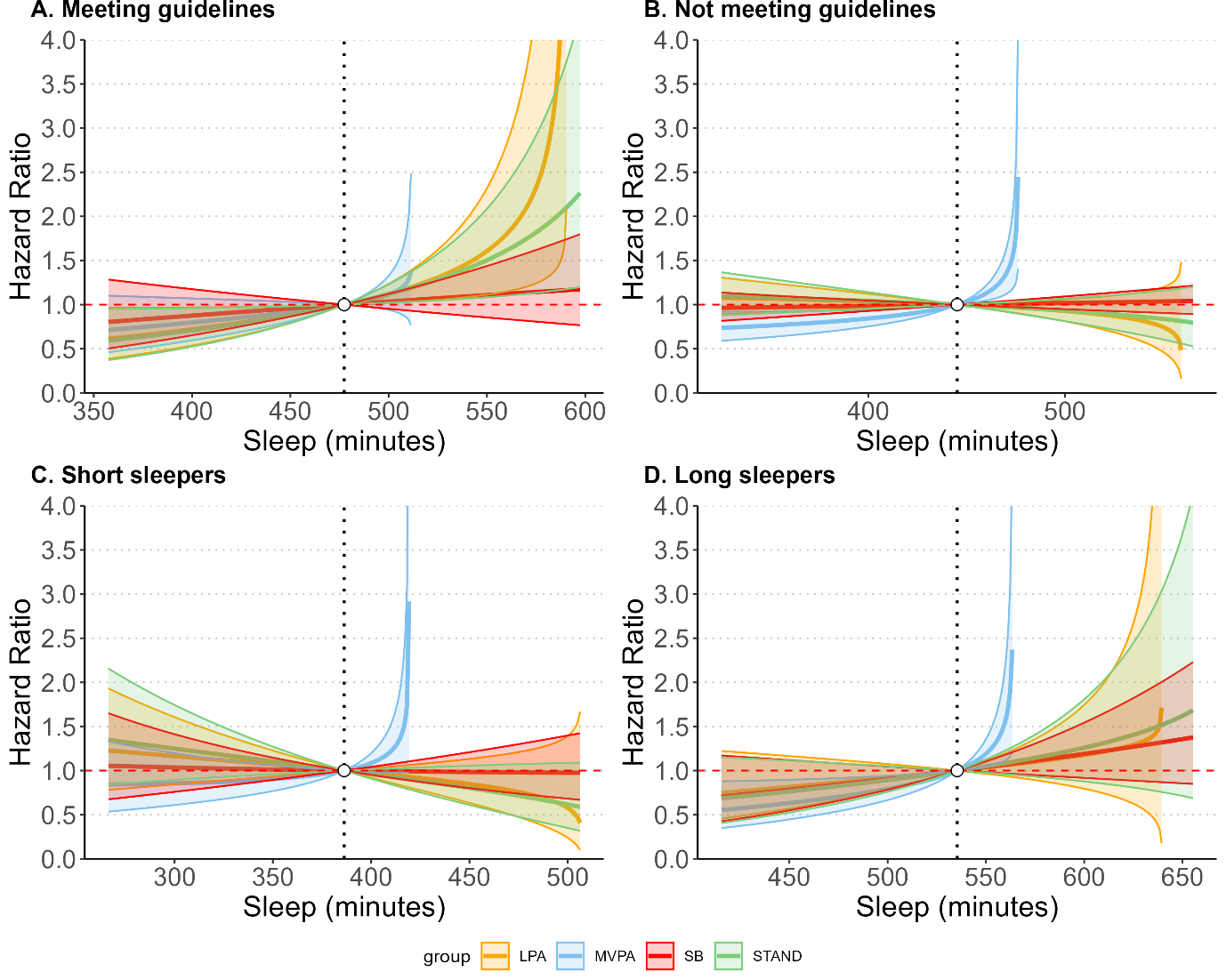

Estimated hazard ratios with 95%CIs in cancer mortality associated with the reallocation of time between sleep and other behaviours across four sleep duration groups: A. meeting sleep duration guidelines (n=28,155; 428 events); B. not meeting guidelines (n=24,046; 549 events); C. short sleep (below guidelines) (n=14,008; 291 events); D. long sleep (above guidelines) (n=10,038; 258 events).

### eFigure 20: Associations of time reallocation between sleep and other movement behaviours with all-cause mortality stratified by sleep regularity excluding 1,120 participants with poor sleep

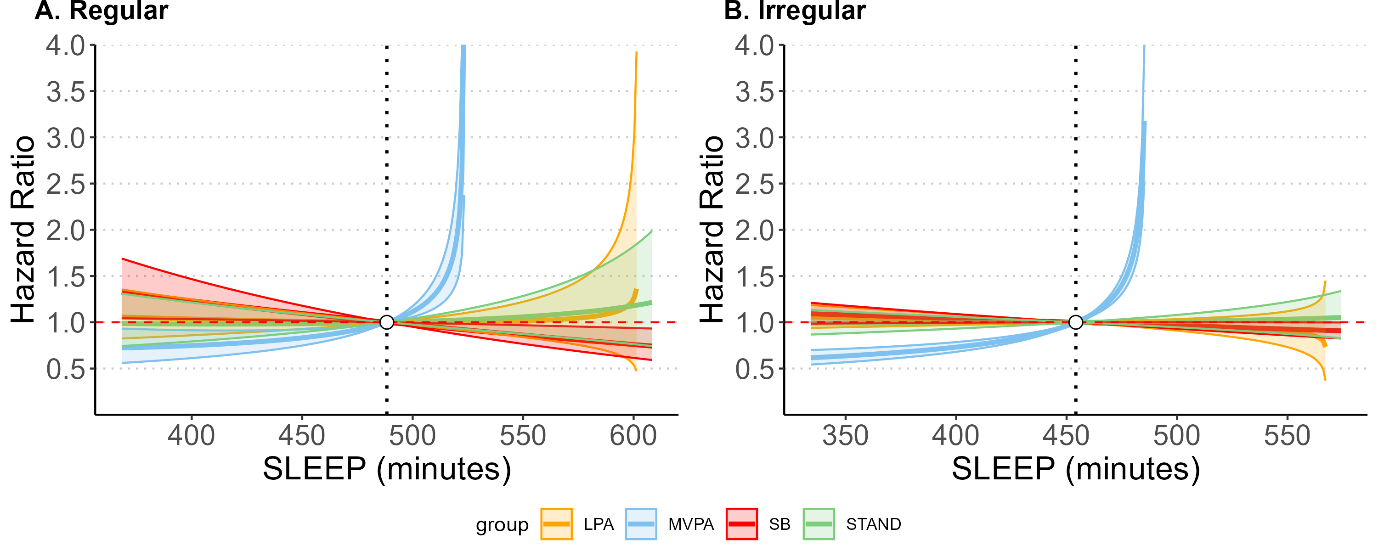

Estimated hazard ratios with 95%CIs in all-cause mortality associated with the reallocation of time between sleep and other behaviours across sleep regularity groups: A. regular sleep (n=14,237; 509 events); B. irregular sleep (n=42,792; 1,648 events).

### eFigure 21: Associations of time reallocation between sleep and other movement behaviours with all-cause mortality stratified by sleep duration adjusting for seasonality

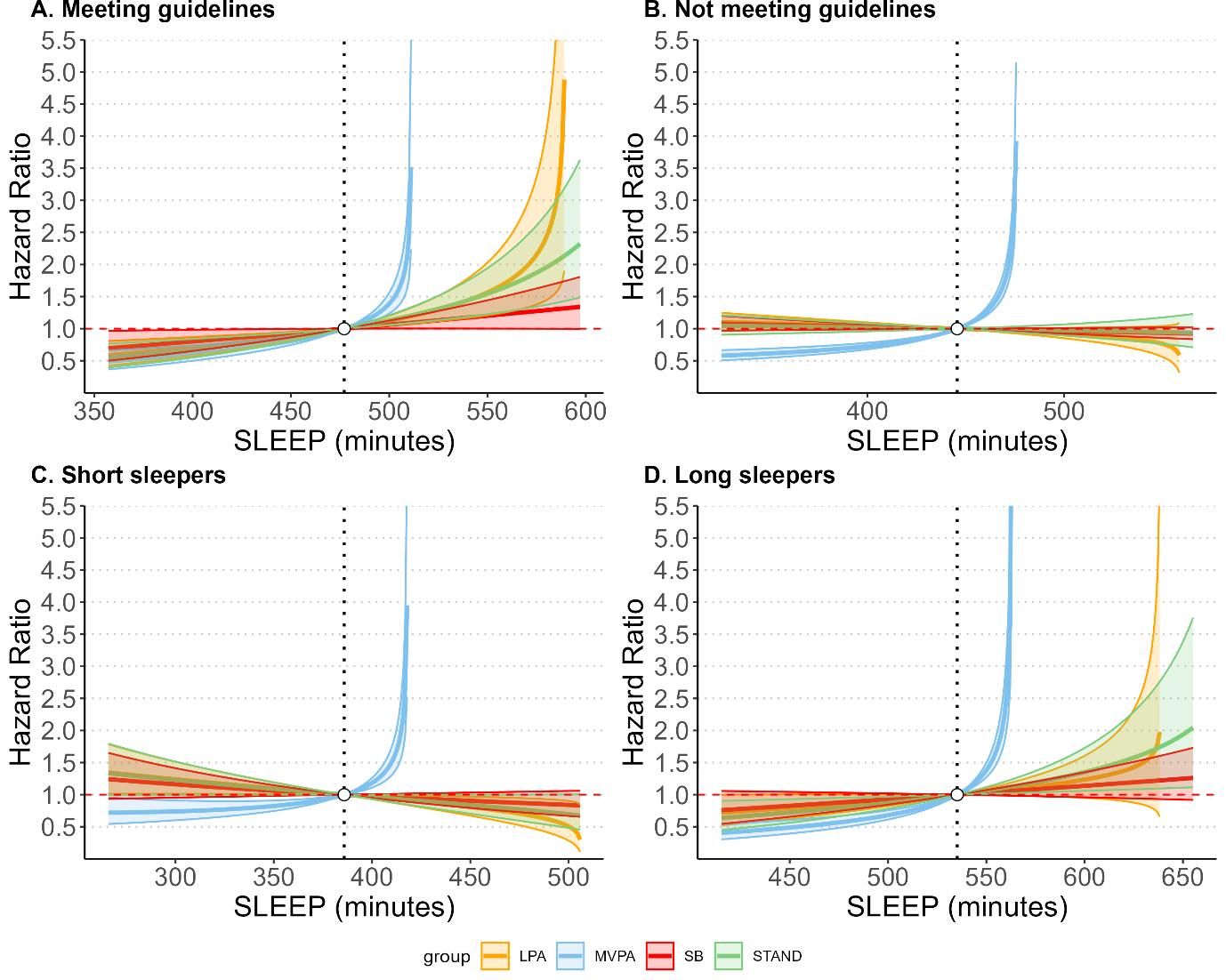

Estimated hazard ratios with 95%CIs in all-cause mortality associated with the reallocation of time between sleep and other behaviours across four sleep duration groups: A. meeting sleep duration guidelines (7–9 hours/day for ages 18–64 and 7–8 hours/day for ages ≥65) (n=30,870; 901 events); B. not meeting guidelines (n=27,279; 1,308 events); C. short sleep (below guidelines) (n=15,739; 711 events); D. long sleep (above guidelines) (n=11,540; 597 events).

### eFigure 22: Associations of time reallocation between sleep and other movement behaviours with CVD mortality stratified by sleep duration adjusting for seasonality

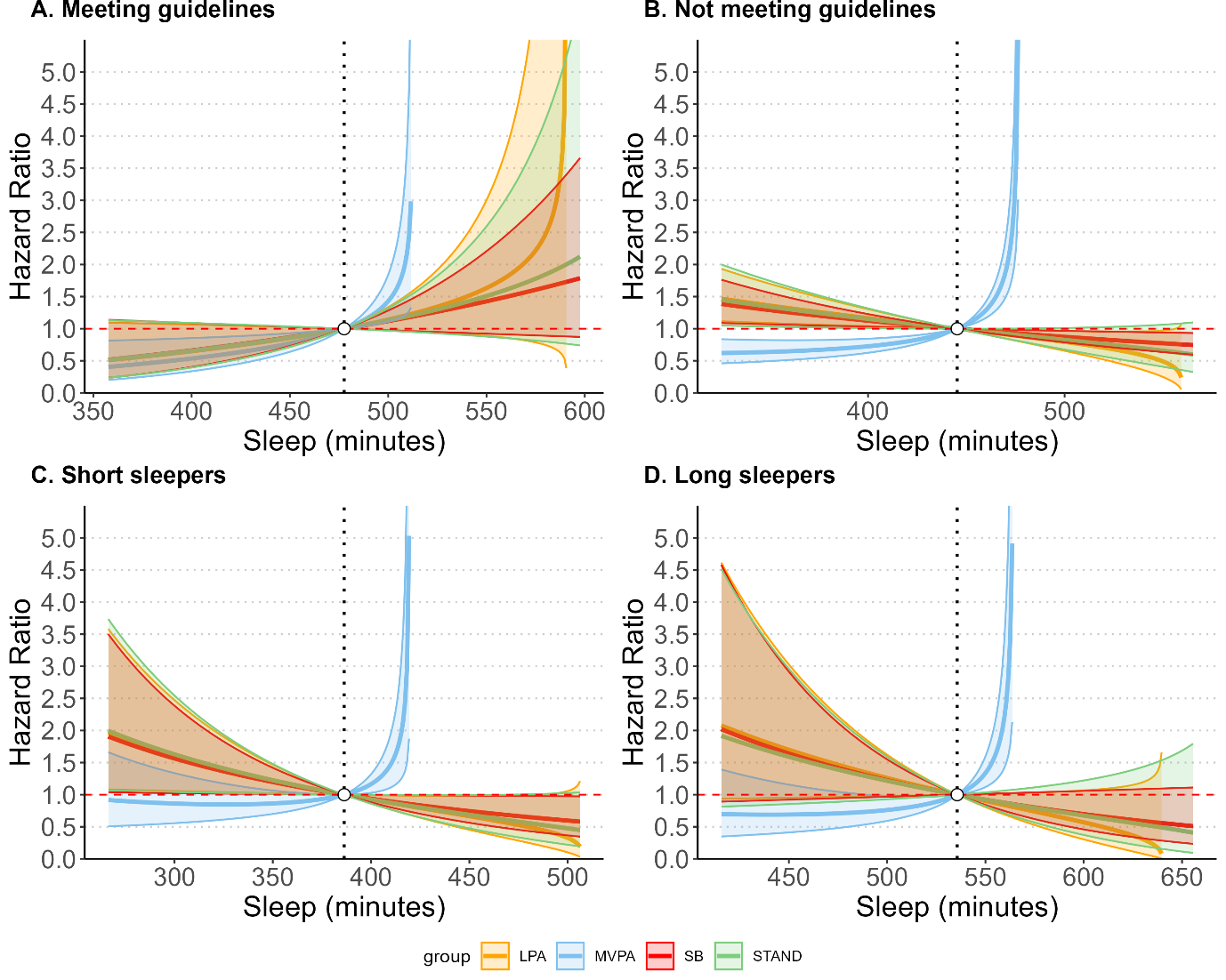

Estimated hazard ratios with 95%CIs in CVD mortality associated with the reallocation of time between sleep and other behaviours across four sleep duration groups: A. meeting sleep duration guidelines (7–9 hours/day for ages 18–64 and 7–8 hours/day for ages ≥65) (n=28,653; 169 events); B. not meeting guidelines (n=24,372; 266 events); C. short sleep (below guidelines) (n=14,184; 156 events); D. long sleep (above guidelines) (n=10,188; 110 events).

### eFigure 23: Associations of time reallocation between sleep and other movement behaviours with cancer mortality stratified by sleep duration adjusting for seasonality

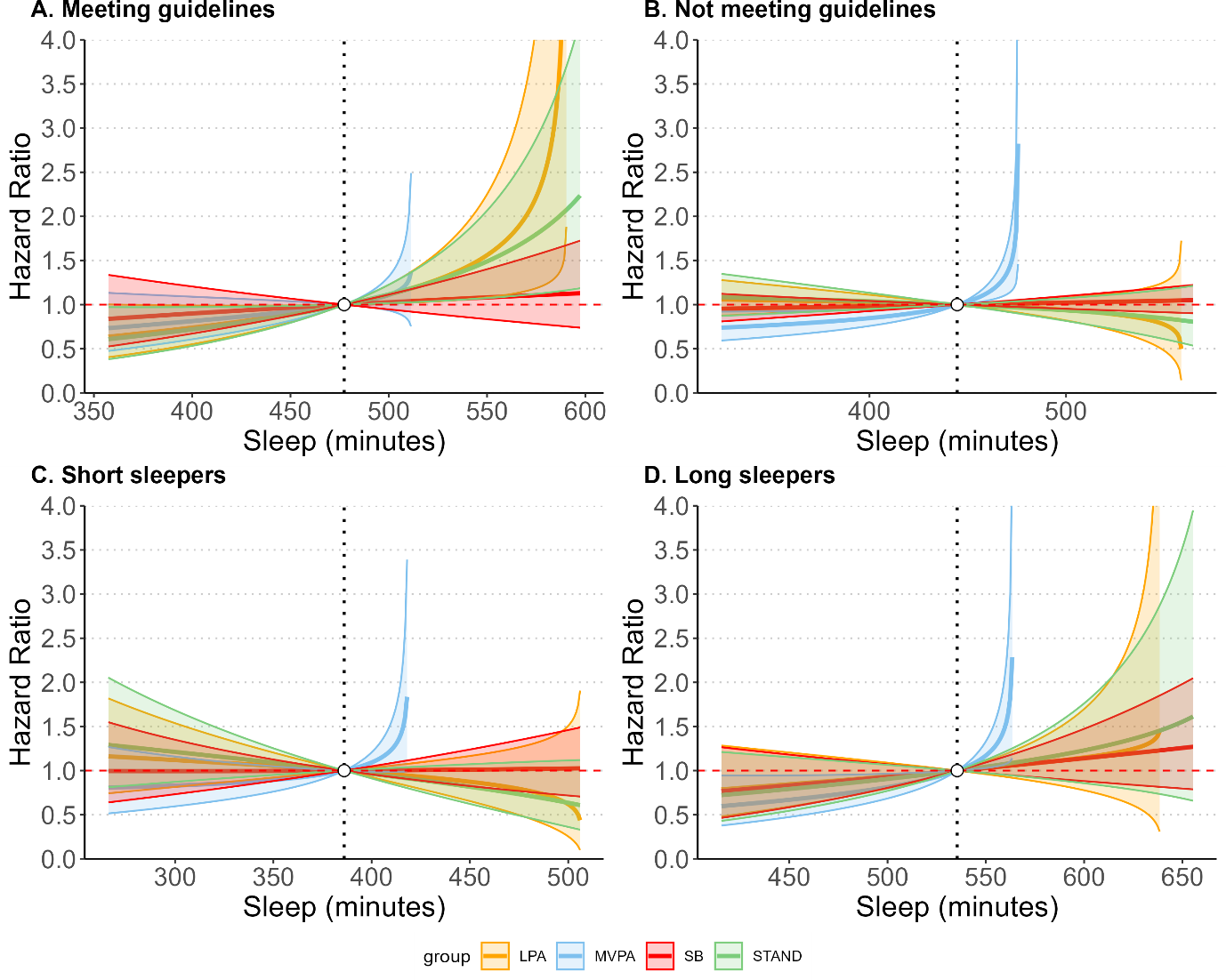

Estimated hazard ratios with 95%CIs in cancer mortality associated with the reallocation of time between sleep and other behaviours across four sleep duration groups: A. meeting sleep duration guidelines (7–9 hours/day for ages 18–64 and 7–8 hours/day for ages ≥65) (n=28,584; 435 events); B. not meeting guidelines (n=24,625; 562 events); C. short sleep (below guidelines) (n=14,415; 298 events); D. long sleep (above guidelines) (n=10,210; 264 events).

### eFigure 24: Associations of time reallocation between sleep and other movement behaviours with all-cause mortality stratified by sleep regularity adjusting for seasonality

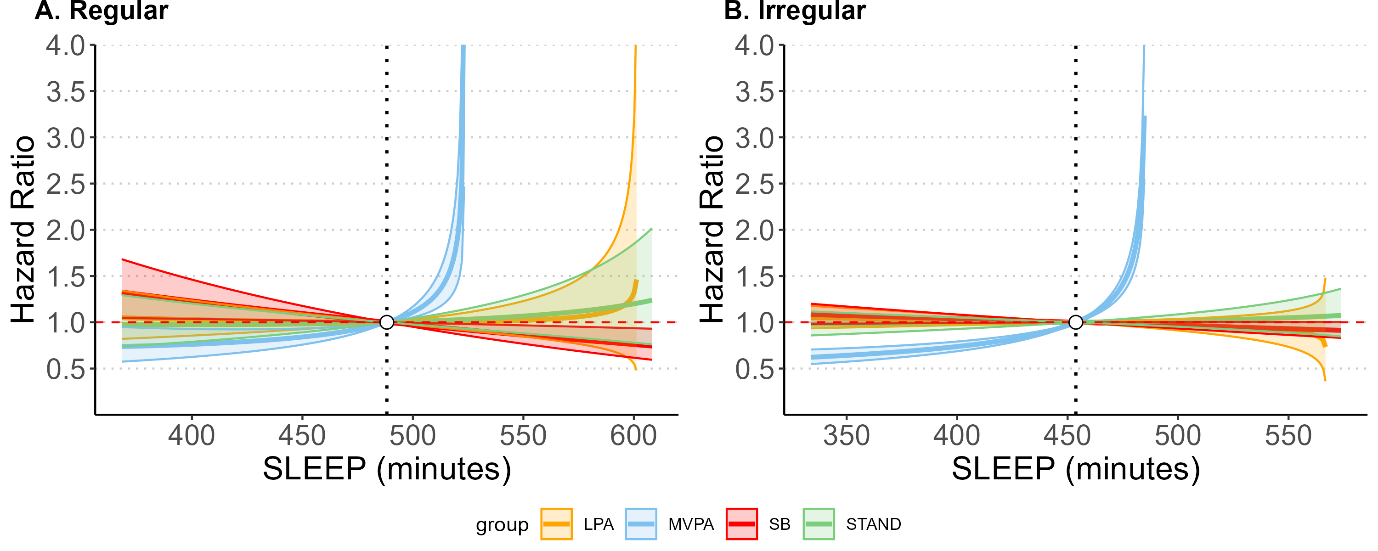

Estimated hazard ratios with 95%CIs in all-cause mortality associated with the reallocation of time between sleep and other behaviours across sleep regularity groups: : A. regular sleep (n=14,507; 517 events); B. irregular sleep (n=43,642; 1,692 events).

### eFigure 25: Associations of time reallocation between sleep and other movement behaviours with all-cause mortality stratified by sleep regularity with stringent classification

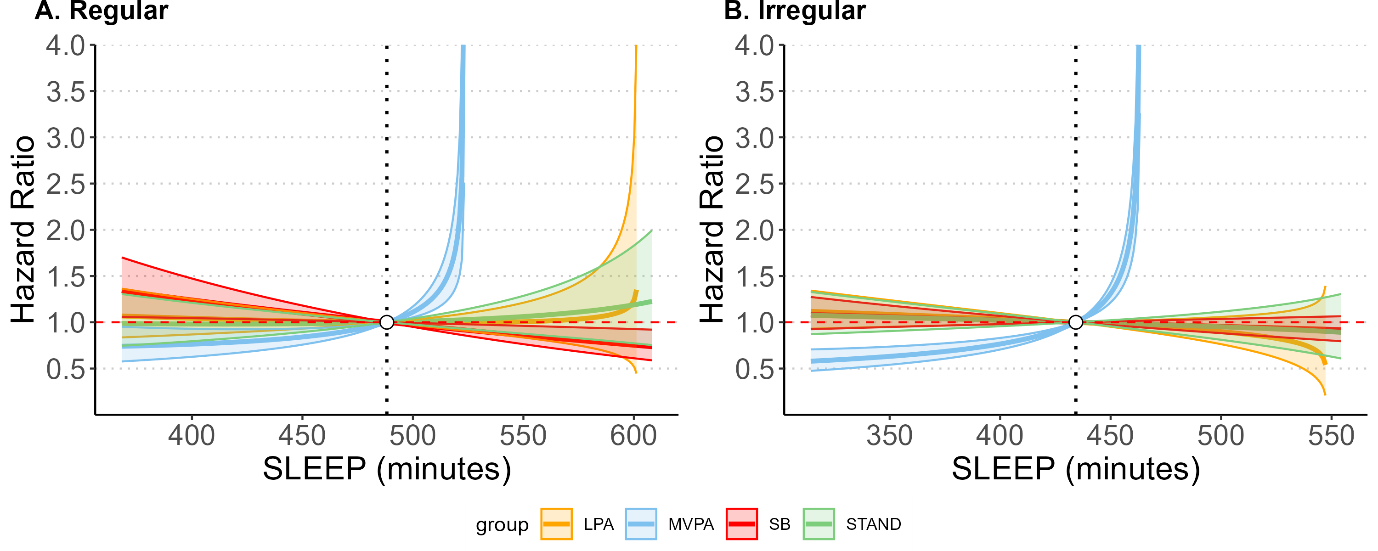

Estimated hazard ratios with 95%CIs in all-cause mortality associated with the reallocation of time between sleep and other behaviours across sleep regularity groups: A. regular sleep (bottom quintile 0-20%; SRI≤70.6) (n=11,597; 430 events); B. irregular sleep (top quintile 80%-100%; SRI>89.0) (n=11,659; 542 events).

### eFigure 26: Associations of time reallocation between sleep and other movement behaviours with all-cause cancer mortality stratified by sleep duration

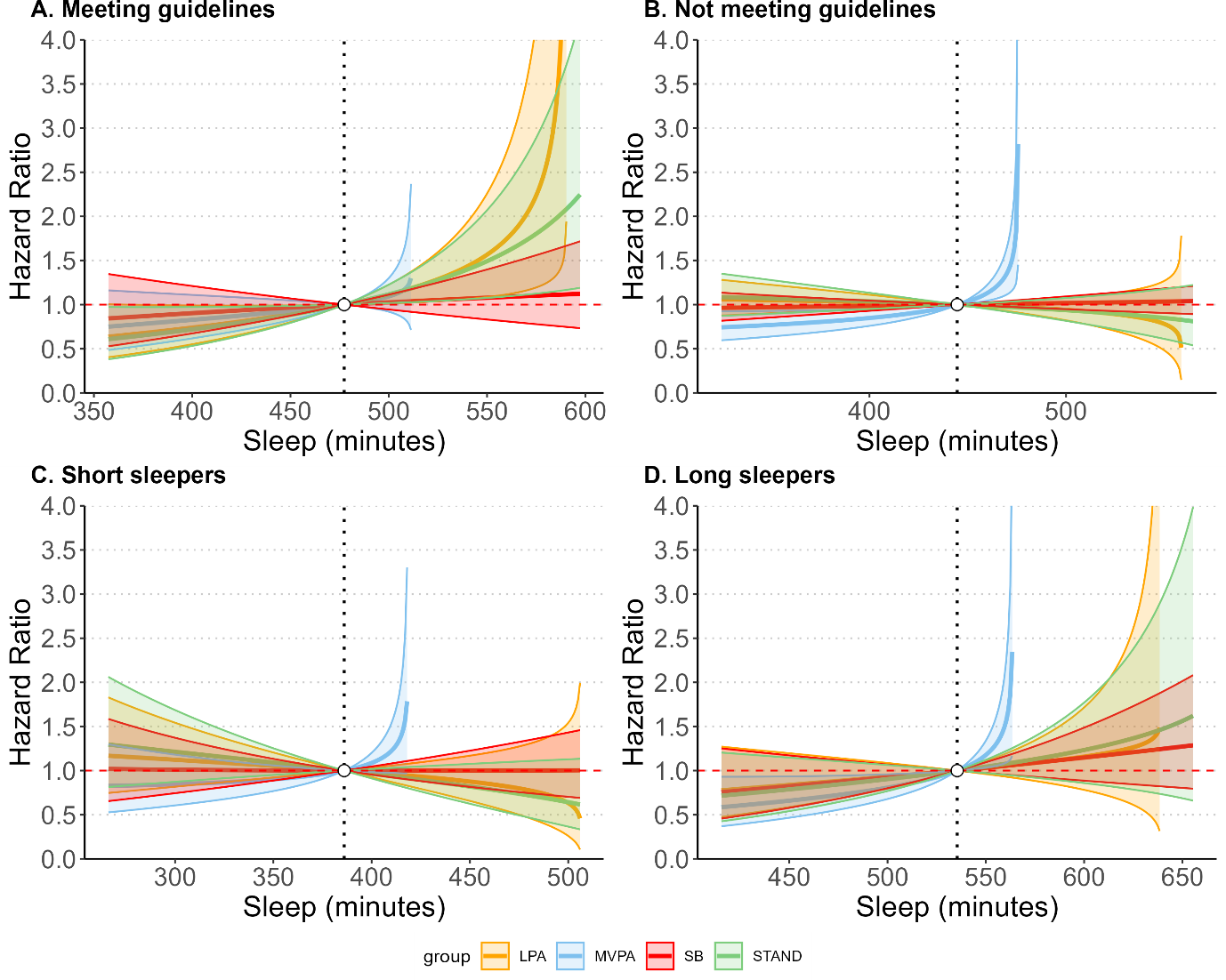

Estimated hazard ratios with 95%CIs in cancer mortality associated with the reallocation of time between sleep and other behaviours across four sleep duration groups: A. meeting sleep duration guidelines (n=28,584; 433 events); B. not meeting guidelines (n=24,625; 555 events); C. short sleep (below guidelines) (n=14,415; 296 events); D. long sleep (above guidelines) (n=10,210; 259 events).

### eFigure 27: Associations of time reallocation between sleep and other movement behaviours with all-cause mortality stratified by sleep duration excluding 13,640 participants with medication use

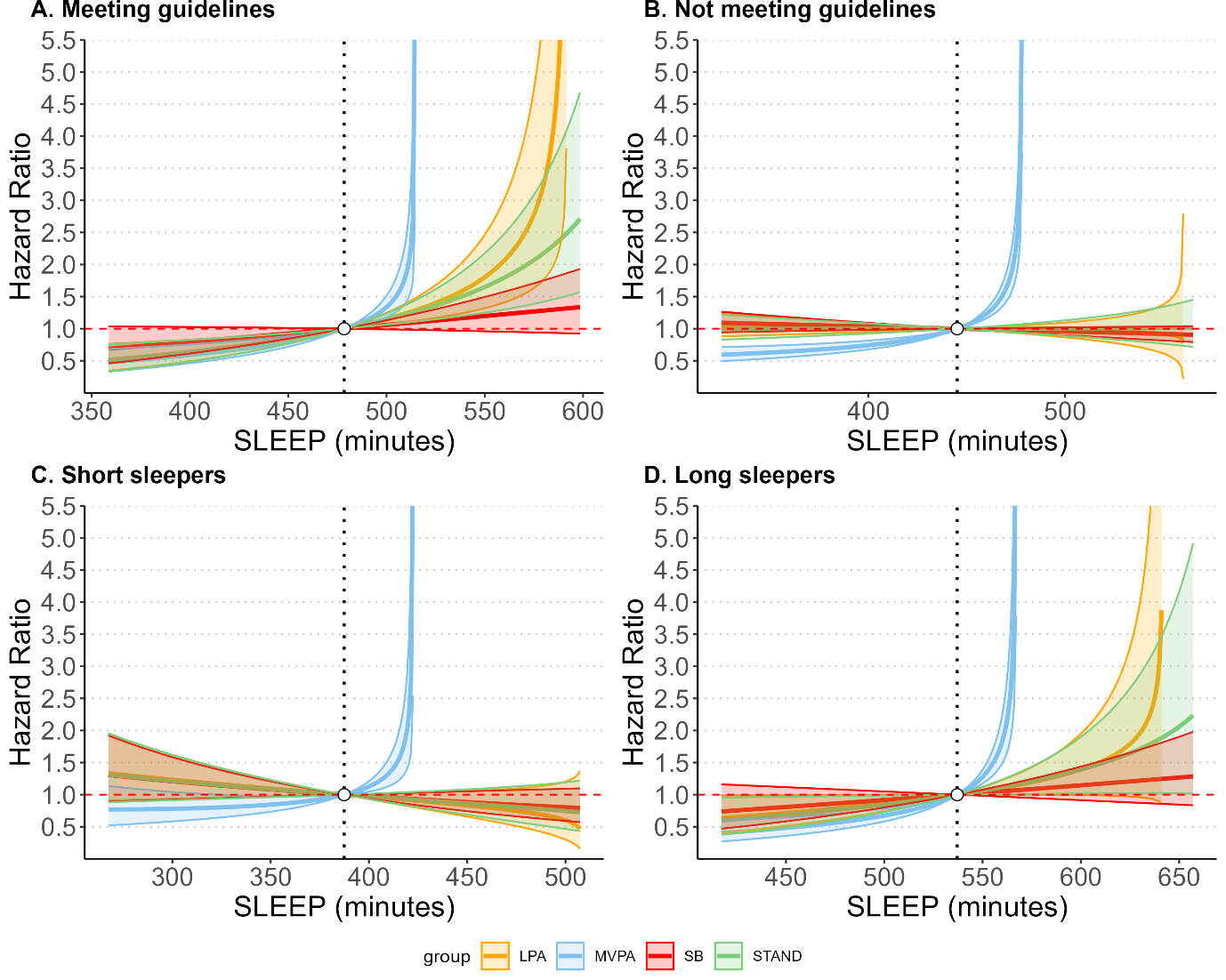

Estimated hazard ratios with 95%CIs in all-cause mortality associated with the reallocation of time between sleep and other behaviours across four sleep duration groups: A. meeting sleep duration guidelines (n=24,938; 550 events); B. not meeting guidelines (n=19,571; 717 events); C. short sleep (below guidelines) (n=11,590; 398 events); D. long sleep (above guidelines) (n=7,981; 328 events).

### eFigure 28: Associations of time reallocation between sleep and other movement behaviours with CVD mortality stratified by sleep duration excluding 13,640 participants with medication use

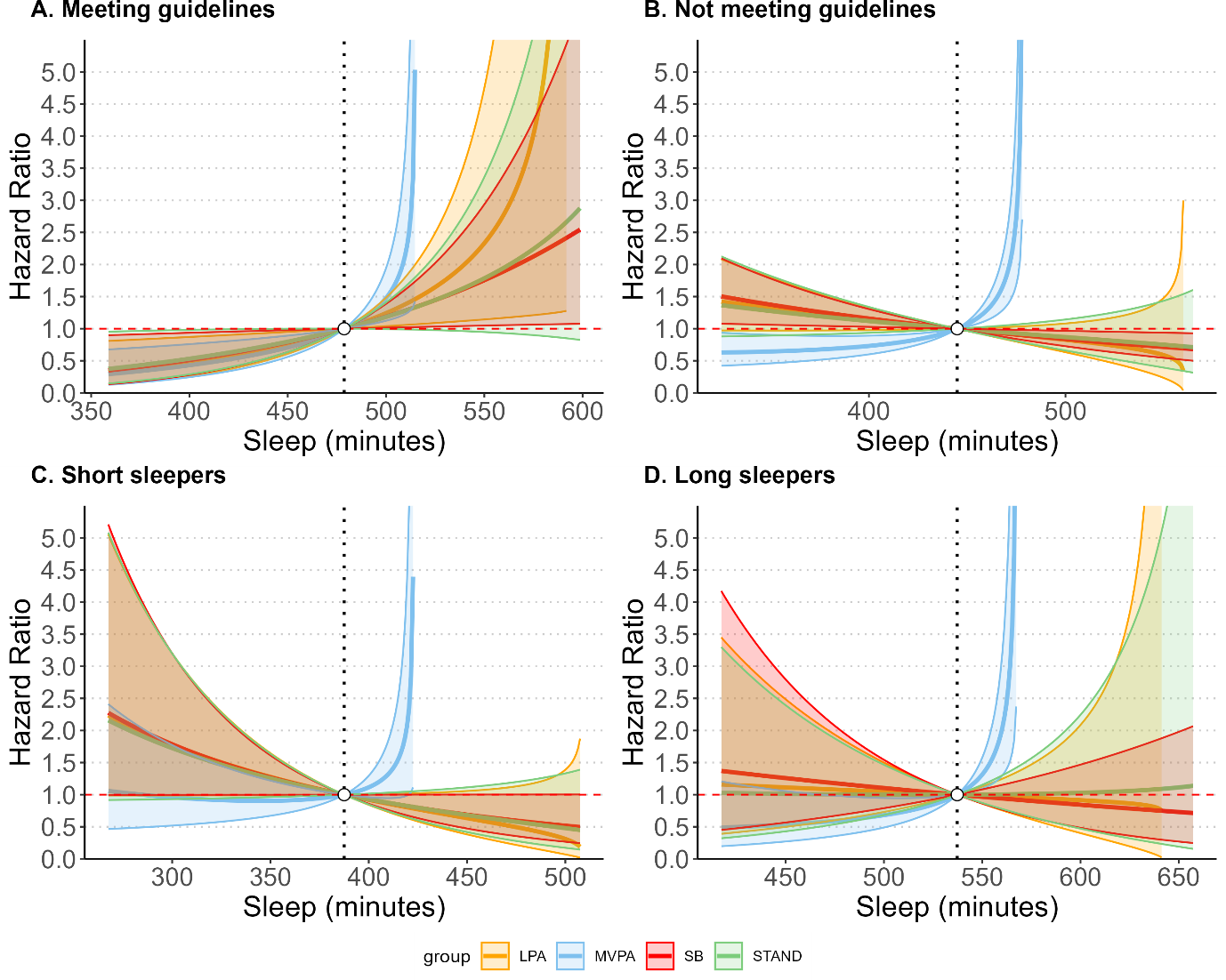

Estimated hazard ratios with 95%CIs in CVD mortality associated with the reallocation of time between sleep and other behaviours across four sleep duration groups: A. meeting sleep duration guidelines (n=23,896; 109 events); B. not meeting guidelines (n=18,485; 140 events); C. short sleep (below guidelines) (n=10,987; 82 events); D. long sleep (above guidelines) (n=7,498; 58 events).

### eFigure 29: Associations of time reallocation between sleep and other movement behaviours with cancer mortality stratified by sleep duration excluding 13,640 participants with medication use

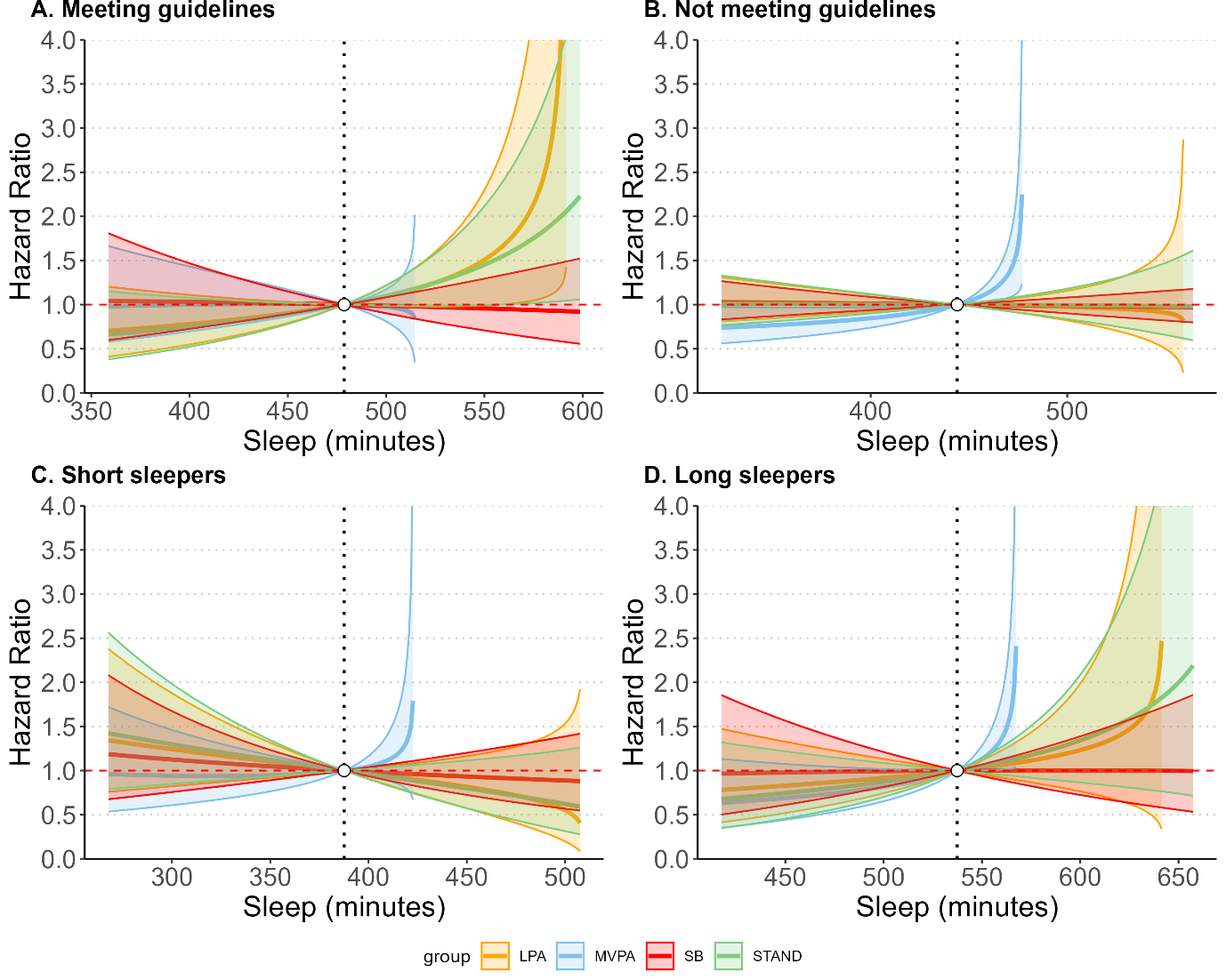

Estimated hazard ratios with 95%CIs in cancer mortality associated with the reallocation of time between sleep and other behaviours across four sleep duration groups: A. meeting sleep duration guidelines (n=23,228; 297 events); B. not meeting guidelines (n=17,833; 347 events); C. short sleep (below guidelines) (n=10,723; 186 events); D. long sleep (above guidelines) (n=7,110; 161 events).

### eFigure 30: Associations of time reallocation between sleep and other movement behaviours with all-cause mortality stratified by sleep regularity excluding 13,640 participants with medication use

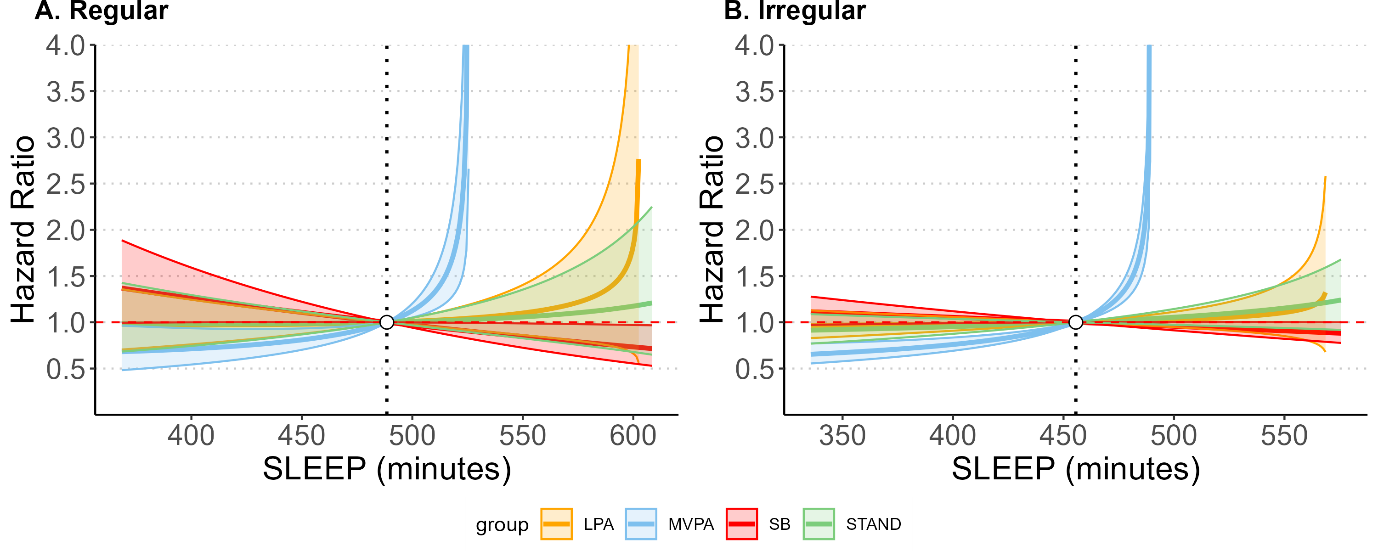

Estimated hazard ratios with 95%CIs in all-cause mortality associated with the reallocation of time between sleep and other behaviours across sleep regularity groups: A. regular sleep (bottom quintile 0-20%; SRI≤70.6) (n=11,084; 297 events); B. irregular sleep (top quintile 80%-100%; SRI>89.0) (n=33,425; 970 events).

### eFigure 31: Associations of time reallocation between sleep and other movement behaviours with all-cause mortality stratified by sleep duration excluding 9,353 participants with prevalent CVD and cancer

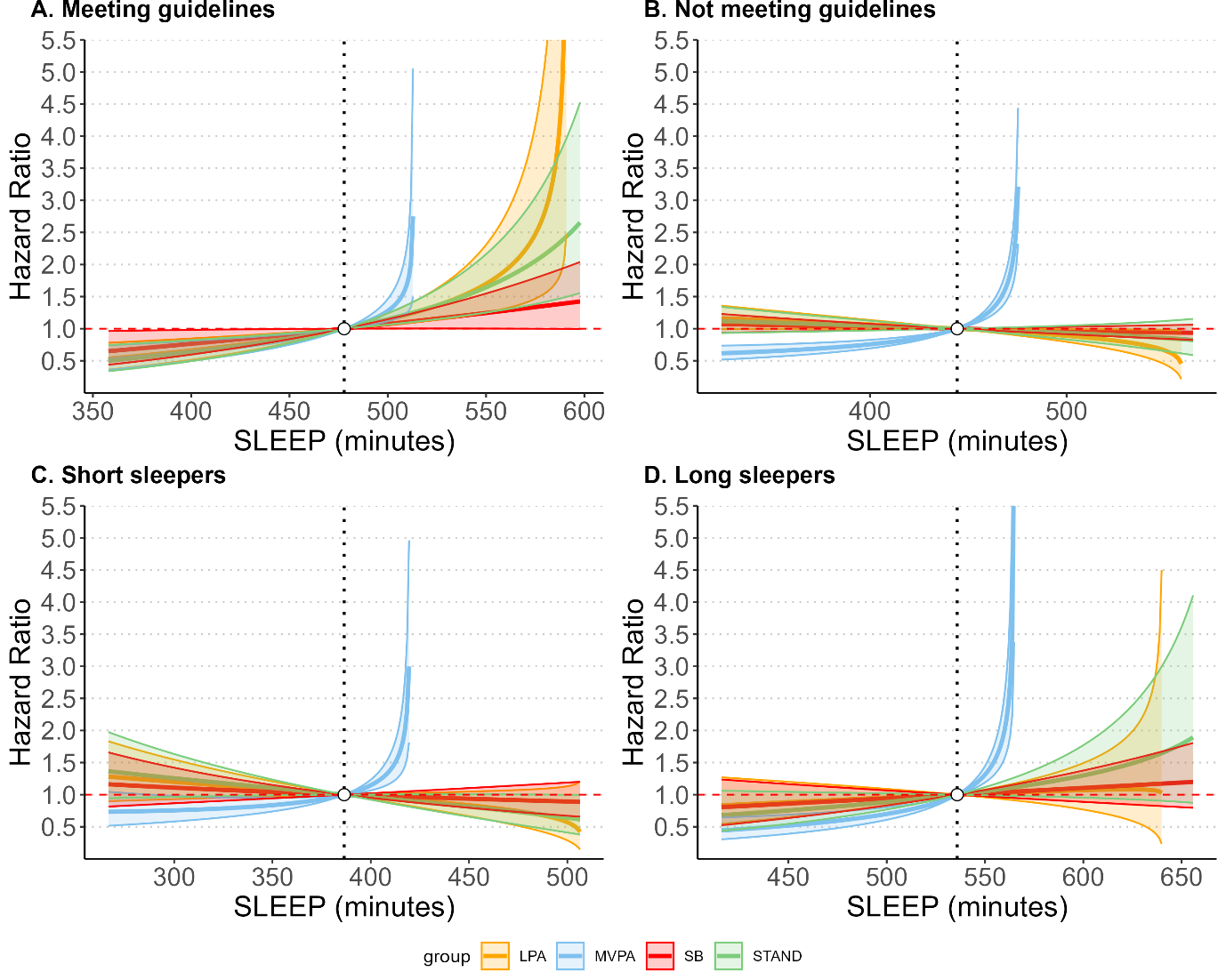

Estimated hazard ratios with 95%CIs in all-cause mortality associated with the reallocation of time between sleep and other behaviours across four sleep duration groups: A. meeting sleep duration guidelines (n=26,611; 605 events); B. not meeting guidelines (n=22,185; 818 events); C. short sleep (below guidelines) (n=13,094; 459 events); D. long sleep (above guidelines) (n=9,091; 359 events).
